# Estimating *Plasmodium falciparum* Parasite Rate using Test Positivity Rate from 2016-2024: Health Management Information Systems in Uganda

**DOI:** 10.64898/2026.02.25.26347098

**Authors:** Jaffer Okiring, John Rek, Austin R. Carter, Juliet N. Nakakawa, Doreen Mbabazi, Thomas Eganyu, Meddy Rutayisire, Catherine M. Sebuguzi, Paul Mbaka, Jimmy Opigo, Dorothy Echodu, David L. Smith, Dianna E.B. Hergott

## Abstract

**Background:** Malaria transmission in Uganda is heterogenous, so the national malaria program needs information about the distribution of malaria to develop appropriate policies. While population-based community surveys estimate *Plasmodium falciparum* parasite rate (*Pf*PR), they are too infrequent and sparse for routine malaria management. Health facility data is routinely collected and covers a large geographic scope, but the data is collected passively, variable in quality, and potentially highly biased. We aimed to triangulate test positivity rate (TPR) from health facility data to survey estimated *Pf*PR data in Uganda to create monthly, high-resolution *Pf*PR estimates.

**Methods:** Using matched health facility and survey data, we fit a multi-level logistic regression model that accounted for clustering at the district and region level, to predict *Pf*PR from TPR. Additional covariates were explored to select a final model that reduced bias while prioritizing its utility for programmatic tasks. Model predictions were validated against observed *Pf*PR and used to generate monthly district-level prevalence estimates from 2016 to 2024. Regional and national level estimates were made by weighting district level estimates by population.

**Results:** The final model included a smoothed TPR term and proportion of severe malaria cases at a district-month level. Predicted *Pf*PR was strongly positively correlated with the observed survey *Pf*PR (Pearson’s rank correlation rho =0.79, p<0.001). National estimates derived from predicted *Pf*PR aligned well with survey estimates from the same time and area.

**Conclusion:** Health Management Information System (HMIS) data, when paired with research data, can be used to estimate malaria prevalence with high spatial and temporal resolution. Estimates can be tested and models can be updated to help malaria programs best leverage facility data. In the context of declining survey frequency, HMIS-based modeling offers a resilient and cost-effective alternative for malaria surveillance and programmatic decision-making in Uganda and similar high-burden settings.

## Introduction

Despite continued investments and innovation in malaria control, the number of malaria cases has remained steady over time. Approximately 282 million cases of malaria occurred in 2024, mostly in sub-Saharan Africa ^1^. The geographical distribution of malaria varies due to a range of factors relating to humans, mosquitoes, parasites, economic development, and malaria control. As control efforts continue to reduce malaria transmission, several countries, including Uganda, are adopting sub-national tailoring (SNT) approaches, as recommended by the WHO ^2^. SNT is a process of data-informed decision making, in which control measures are adapted based on transmission levels. At the same time, national malaria programs are tasked with managing malaria in real time. For National Malaria Control Programs (NMCPs) to properly plan and implement SNT and manage malaria, they need access to frequent, high-resolution, up-to-date, accurate measures of malaria transmission.

Research metrics commonly used to measure malaria transmission intensity include *Plasmodium falciparum* parasite rate (*Pf*PR, the proportion of the population infected), and less commonly, the entomological inoculation rate (EIR, number of infectious bites per person per time). Surveillance metrics, derived mostly from data collected at health facilities, include clinical incidence (the number of confirmed malaria cases/100,000 population) ^3,4^, and the malaria test positivity rate. While valuable, each measure has limitations that reduce its usability for frequent planning and control.

Over the past few decades, malaria planning has been based on the *Pf*PR. *Pf*PR estimates predominantly come through national, population-based malaria surveys such as the Malaria Indicator Survey (MIS) and Demographic and Health Surveys (DHS), but data collected as part of malaria research is an important alternative source. While these surveys are well-designed ^5–7^, they are costly and thus conducted infrequently. Additionally, MIS and DHS estimates are usually restricted to children under five years of age, providing limited insight into transmission in older populations^8^. Sophisticated Bayesian geostatistical methods address some of these shortcomings by using environmental covariates to help fill gaps in space, time, and age. These methods produce high resolution maps of malaria *Pf*PR for national malaria programs ^9–11^, but the utility of these maps is ultimately limited by availability of data.

Given the need for timely and extensive data to manage malaria, malaria programs must build information systems that make the most of health facility data. Two metrics commonly reported from a health management information system (HMIS) for malaria are the total tests performed and the number that were positive, which allows for the calculation of the test positivity rate (TPR). Several countries maintain a HMIS, where routine information is collected from health facilities throughout the country on a weekly or monthly basis. As the data coming in is from passive surveillance, there are questions about the accuracy and quality of the underlying data. Data from passive streams can only be validated through methods that pair surveillance metrics with prospective studies and active study designs. In other words, malaria programs must develop methods to understand what information about malaria in populations can be accurately inferred from facility data. Importantly, Bayesian geostatistical methods have been updated to use facility data for long-term planning cycles, but there is a need to improve the use of malaria surveillance in routine management of malaria^12^.

There is no *a priori* basis for using these surveillance metrics as a quantitative basis for understanding malaria in local populations. Several studies have sought to model the relationship between TPR and clinical incidence ^13–15^. While valuable, the models are often complex, and rely on having facility catchment areas, which limits their usefulness to a small proportion of the data. An alternative approach is to relate TPR to *Pf*PR, as both of these measures are not dependent on the number of patients coming to a health facility ^13,16^ or a known catchment area. However, TPR is an inherently biased measure, influenced by changes in diagnostic testing methods and propensity to test, healthcare-seeking behaviour, and the incidence of non-malarial febrile illnesses ^17^. While TPR has known biases, it is widely available and constantly updated, and could be valuable to inform malaria programs if it could be used to estimate *Pf*PR more frequently. A few studies have sought to quantify this relationship, using cross-sectional data gathered and showing a non-linear relationship ^18,19^.

In this study, we developed a statistical model to translate TPR into *Pf*PR for all districts in Uganda, using HMIS data and *Pf*PR estimates from three nationally representative surveys. Our goal was to create a model that is both accurate and easily updateable, enabling NMCPs to monitor malaria burden in near real-time and support sub-national tailoring of interventions. Given concerns about bias, we sought covariates that could help with the bias. In selecting covariates for the model, the rule was to prefer covariates that could improve the predictive accuracy in a timely way, so we would avoid using any covariates that would complicate the provision of timely data unless they produced a large improvement in the predictive accuracy.

## Methods

### Malaria in Uganda

Malaria transmission in Uganda occurs throughout the year and varies across the country, with a potential of epidemics ^20,21^. Of note, regions such as West Nile, Acholi, Lango, Karamoja, Teso, Bukedi, and Busoga experience high malaria transmission while the South-central, North-central, Kampala, Ankole, Tooro, Bunyoro, and Kigezi regions generally have lower transmission intensities ^22,23^. Malaria transmission levels have been reported to peak during and after the two rainy seasons, specifically from March to May and from September to November ^24–26^. To combat malaria, Uganda conducts universal distribution of long-lasting insecticidal nets (LLINS) every three years since 2013. Additionally, higher transmission areas have received indoor residual spraying (IRS) ^27^ and seasonal chemoprophylaxis (SMC) ^28,29^.

Uganda’s Ministry of Health has maintained a HMIS, hosted on the DHIS2 platform, since 2015. Data is submitted in aggregate both weekly and monthly, and over time, the quality of HMIS data has gradually improved. Approximately 4000 health facilities report into HMIS.

### Data sources

#### *Pf*PR Data

*PfP*R data for model fitting was obtained from three representative surveys: the 2016 DHS, 2018 MIS, and LLIN Evaluation in Uganda Project (LLINEUP) 2017-2019 for Uganda. Individual level data from each study, including age, sex, cluster, district, malaria test performed, malaria test result, and month and year of testing were extracted. Information on the MIS and DHS studies has been published previously ^30,31^. LLINEUP was a cluster-randomized trial of LLINs in 104 of 221 health sub-districts, located in 48 districts in Uganda ^32–34^. Children aged 2-10 years enrolled were tested throughout the study. Only data from those under 5 were included in this analysis.

### Test Positivity Rate (TPR) data

Data on all-age confirmed malaria cases, suspected cases, tests performed (microscopy or RDT), outpatient visits, and inpatient malaria cases were extracted from all health facilities reporting to HMIS between 2015 and 2024. Monthly test positivity rate (TPR) was calculated as the proportion of all tests performed that were positive for malaria for each district, providing estimates for each district-month.

### Exploratory Covariates

Additional covariates were explored to see if they improved model fit, including temperature, rainfall, population density, and median walk time to the nearest facility. Temperature and rainfall data were obtained from the United States Geological Survey (USGS) Famine Early Warning Systems Network (FEWS NET) Data Portal (https://earlywarning.usgs.gov/fews/software-tools/). This portal links to different datasets like Climate Hazards Group InfraRed Precipitation with Station data (CHIRPS) rainfall and Land Surface Temperature (LST) ^35,36^. CHIRPS rainfall and Land Surface Temperature were requested for 2015 to 2024, by month. Using district shapefiles, average monthly rainfall and land surface temperature were extracted for each district.

Population density for each district was obtained using estimates published in the 2024 household census from UBOS ^37^. Median walk time to the nearest facility was calculated utilizing the geo-located points of health facilities reporting to the HMIS and the walking friction surface created by Malaria Atlas Project (MAP)^38^. The distance to the nearest health facility for all populated 1kmx1km pixels was calculated, and then district average was determined by taking a population weighted median of all pixel estimates.

### Data Matching and Preprocessing

Prior to calculating variables from HMIS data, we ran an imputation and outliering algorithm on the raw facility data to remove implausible values and impute missing values for tests, confirmed cases, suspected malaria, and outpatient visits (OPD). For TPR calculations, only facilities who reported data for >50% of the time period, and those that had information on confirmed cases and tests, were included in district level calculations. After imputation, if tests were not reported, or tests were less than confirmed cases, suspected malaria was used as a proxy. The proportion of cases in a district that were severe was calculated by month as the number of inpatient-malaria divided by all confirmed malaria cases, using data from all reporting facilities.

All data were temporally and spatially aligned by matching on district, month, and year. Temporal smoothing techniques were applied to the TPR data to reduce noise and improve model stability. Covariates with limits beyond 0 and 1 were scaled prior to model fitting.

### Model selection

Using our paired data, we fit a mixed-effects logistic regression model of under-five *Pf*PR to 180-day smoothed all ages TPR data (now referred to as TPR), with random effects for district and region. Fit *Pf*PR, estimated from individual level survey data for children under 5, was a binary variable coded as 0 and 1. To determine if additional covariates improved model fit, they were added as fixed effects into the model, first singularly. Any covariates with a significant association in the singular model were then added in all possible combinations. Model selection was based on the Akaike’s information criteria (AIC), while also balancing availability of covariates to be continuously updated for prediction. Model predicted *Pf*PR to measured *Pf*PR was visualized with a scatter plot, and the correlation was estimated using Pearson’s correlation coefficient.

### Prediction of PfPR for each district

After selecting the best fit model from our matched dataset, we estimated monthly *Pf*PR from TPR for all districts from 2016 to 2024. District level TPR estimates were created from HMIS data for all available months as described above and smoothed. Confidence estimates for predictions were estimated through bootstrapping with *BootMer* (R version 4.3.2), performing 1000 simulations. Monthly estimates at higher administrative levels (region and national) were estimated by taking a population weighted mean of the district level *Pf*PR estimates. Yearly averages were calculated by taking the mean of the monthly averages at the appropriate administrative level. To compare to survey level national estimates, equivalent *Pf*PR predictions were obtained by averaging data from HMIS restricted to those from the same districts and months as the survey. An estimate for a 2024 national estimate comparable to survey data was estimated using data from HMIS for all districts for November and December.

## Results

Individual level survey data on malaria positivity was available for 45,396 children under 5 across the three surveys, collected between June 2016 and September 2019. In both MIS and DHS, enumeration area was defined as a cluster while in LLINEUP, health sub district was taken as a cluster. MIS had 340 clusters, DHS had 692, and LLINEUP had 104 clusters. Approximately equal number of males and females were tested in all surveys. MIS and LLINEUP relied on microscopy, while DHS used RDT. Median observed *Pf*PR ranged from 10.4% in LLINEUP to 23.8% in DHS. The median *Pf*PR surveyed was highest among those enrolled in DHS compared to those in MIS and LLINEUP (Table 1).

**Table 1.** Descriptive information for survey data used to fit model. Data comes from the 2016 Demographic Health Survey (DHS), 2018 Malaria Indicator Survey (MIS) and MIS 2018, DHS 2016, and the LLIN Evaluation in Uganda Project (LLINEUP), conducted between 2017 and 2019.

| Characteristics | Surveys |  |  |
| --- | --- | --- | --- |
|  | DHS | MIS | LLINEUP |
| Period of survey | June 2016 - Dec 2016 | Dec 2018 - Jan 2019 | Mar 2017 - Sept 2019 |
| Number of clusters | 692 | 340 | 104 |
| Number of districts | 112 | 123 | 48 |
| Total children under 5 tested | 5500 | 7780 | 13778 |
| Test used | RDT | microscopy | microscopy |
| Number of females (%) | 2728 (49.6) | 3831 (49.2) | 6999 (50.1) |
| Median age (IQR) | 3 (1-4) | 2 (1-3) | 3 (2-4) |
| Median observed *PfPR* (IQR) | 23.8% (9.7%-50.2%) | 11.1% (3.7%-22.5%) | 10.4% (3.7%-21.6%) |

MIS -Malaria Indicator Survey DHS-Demographic and Health Survey LLINEUP-LLIN Evaluation in Uganda Project

Data from more than 3000 health facilities in the HMIS were extracted for district-months with survey data. Table 2 presents a summary of the paired HMIS dataset that was used for model fitting. Unweighted median TPR ranged from 34.0% in 2017 to 46.4% in 2016.

**Table 2.** Description of data utilized from the Health Management Information System (HMIS) that was paired with surveys for model fitting.

| Characteristics | Year |  |  |  |
| --- | --- | --- | --- | --- |
|  | 2016 | 2017 | 2018 | 2019 |
| Median total health facilities reporting per month | 3348 | 1470 | 2926 | 3039 |
| Districts reporting data | 112 | 48 | 81 | 99 |
| Total tested | 2,905,943 | 1,788,793 | 2,207,744 | 1,334,090 |
| Total confirmed malaria | 1,223,864 | 871,050 | 886,045 | 521,697 |
| Total cases of severe malaria | 87,046 | 51,869 | 51,372 | 30,809 |
| Median observed TPR (IQR) | 46.4% (35.9%-55.1%) | 34.0% (18.0%-46.6%) | 44.4% (34.4%-56.1%) | 35.3% (23.7%-48.1%) |

**Table 3.** Comparison of the national level survey estimated *Pf*PR estimates and 95% confidence interval (CI) by year, and model predicted national estimates. For 2024, confidence intervals were not available for the survey measure estimate.

| Survey | Survey Measured Estimate of *PfPR* (95% CI) | Model Predicted Estimate of *PfPR* (95% CI) |
| --- | --- | --- |
| DHS 2016 | 30.7 % (27.8, 33.5%) | 17.1% (4.65, 38.5%) |
| LLINEUP 2017 | 20.0% (16.8%, 23.3%) | 19.9% (4.57, 47.9%) |
| LLINEUP 2018 | 10.8% (8.05, 13.6%) | 14.0% (2.9%, 36.4%) |
| MIS 2018 | 9.2% (7.3%, 11.0%) | 13.5% (3.25, 33.0%) |
| LLINEUP 2019 | 5.0% (2.5%, 7.5%) | 9.5% (1.8, 28.5%) |
| MIS 2024 | 21.1% | 11.7% (3.3, 29.4%) |

### Performance of the model

There was a moderate positive correlation between TPR and *Pf*PR in the training data (rho=0.57, p<0.0001). The best fit model based on AIC value included a 180-day smoothed TPR term, a term for the proportion of cases that were severe for each district-month, and temperature and rain values for each district-month (see Supplemental Table 1). However, given the added complexity of continually updating a model with external temperature and rain variables, we compared predicted fits of our best model to one that just included TPR and proportion severe, both which are derived solely from the HMIS data. Correlation between observed and predicted *Pf*PR was high for both the best model (R=0.81, p<0.0001), and the less complex model (R=0.79, p<0.0001, **Figure 1A**), and differences in predictions for additional data were minimal (see Supplemental Figure 1). As such, the model with smoothed 180-day TPR and proportion severe was selected as our final model. Correlation between observed TPR and predicted *Pf*PR was high (R=0.74, p<0.0001, **Figure 1B**), with greater variation between TPR and *Pf*PR observed at higher levels of TPR. Median observed *Pf*PR for a district-month across all surveys was 10.7% [IQR: 0, 28%], while median predicted *Pf*PR for a district-month using HMIS data was 13.3% [IQR: 4, 29%].

**Figure 1.**
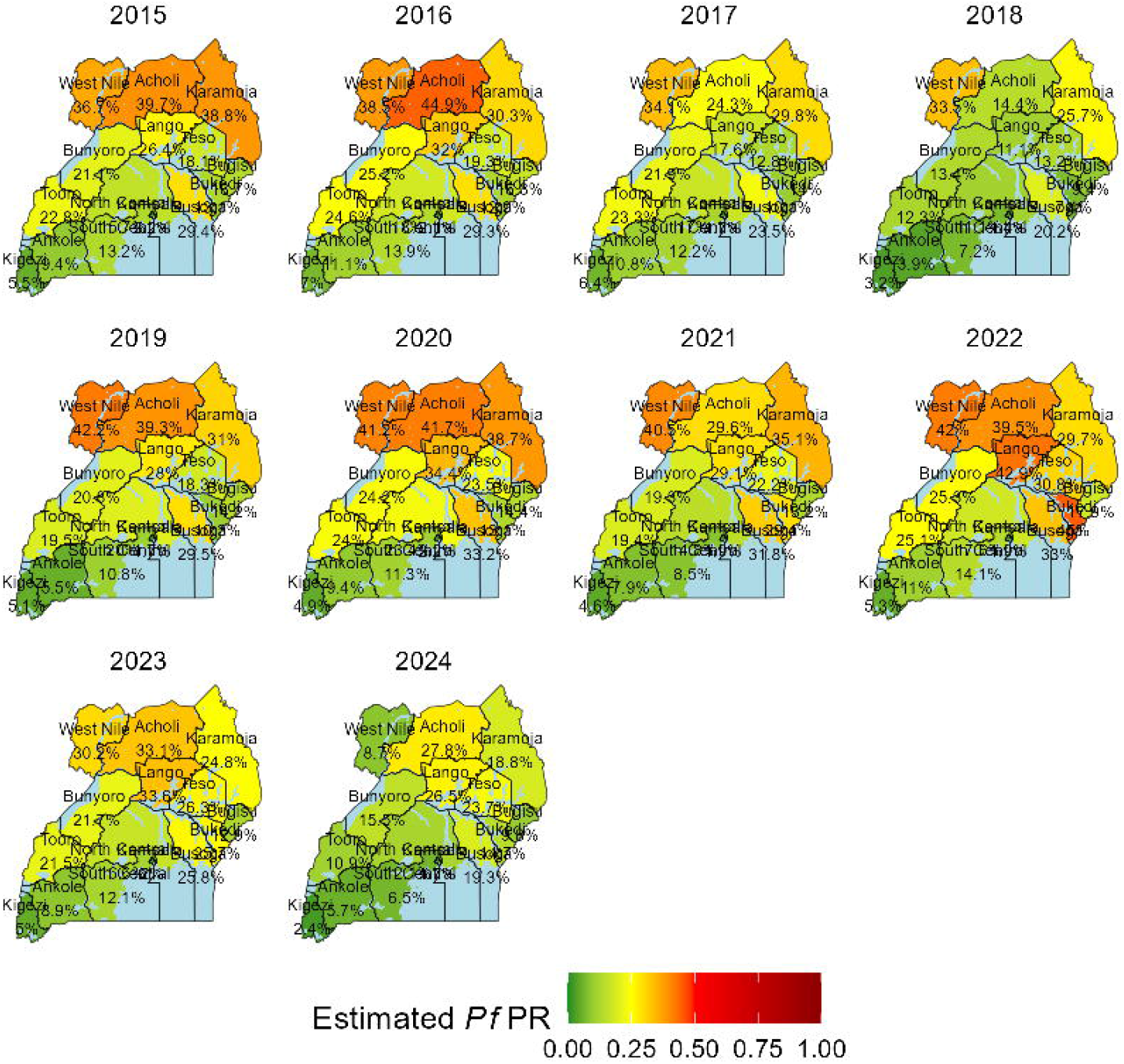
A) Observed *Pf*PR vs predicted *Pf*PR for district-months in the period of 2016-2019 for matched district training data. B) Observed test positivity rate (TPR) vs. predicted *Pf*PR for district-months in the period of 2016-2019 for matched district training data. The size of the dot represents that proportion of all cases that were severe in the district for that month. Dashed line is the linear regression fit of the data, with the Pearson’s correlation coefficient (R) displayed.

### Temporal trends in predicted *Pf*PR at district level

We used our selected model to predict monthly *Pf*PR values for every district from January 2016 to December 2024 using data pulled from the HMIS. **Figure 2** presents the monthly time series of predicted *Pf*PR and survey estimated *Pf*PR for four representative districts with varying underlying levels of endemicity. For all districts, the predicted estimates fall within the range of the survey estimates. Temporal trends that are not apparent with survey data are observed with the predicted *Pf*PR time series. Pader District, a region with consistently high prevalence, demonstrates seasonal peaks in prevalence corresponding to rainy seasons. Additionally, a large dip in prevalence in 2018 is observed, which is consistent with regional trends during this time, possibly due to an IRS round conducted in 2017. Kanungu District, in the low-prevalence region of Kigezi in which no IRS has been done, shows smaller seasonal variation. Yumbe District, in West Nile region, shows that prevalence was high through 2023, when a large decrease was observed as a response to increased vector control methods that began in 2022. Lira District, in the higher prevalence Lango region, shows increasing prevalence around 2020. In this district, IRS efforts were stopped in 2022. Other temporal trends in predicted *Pf*PR among all the districts in Uganda are provided as a supplementary file (Supplementary Figure 2).

**Figure 2.**
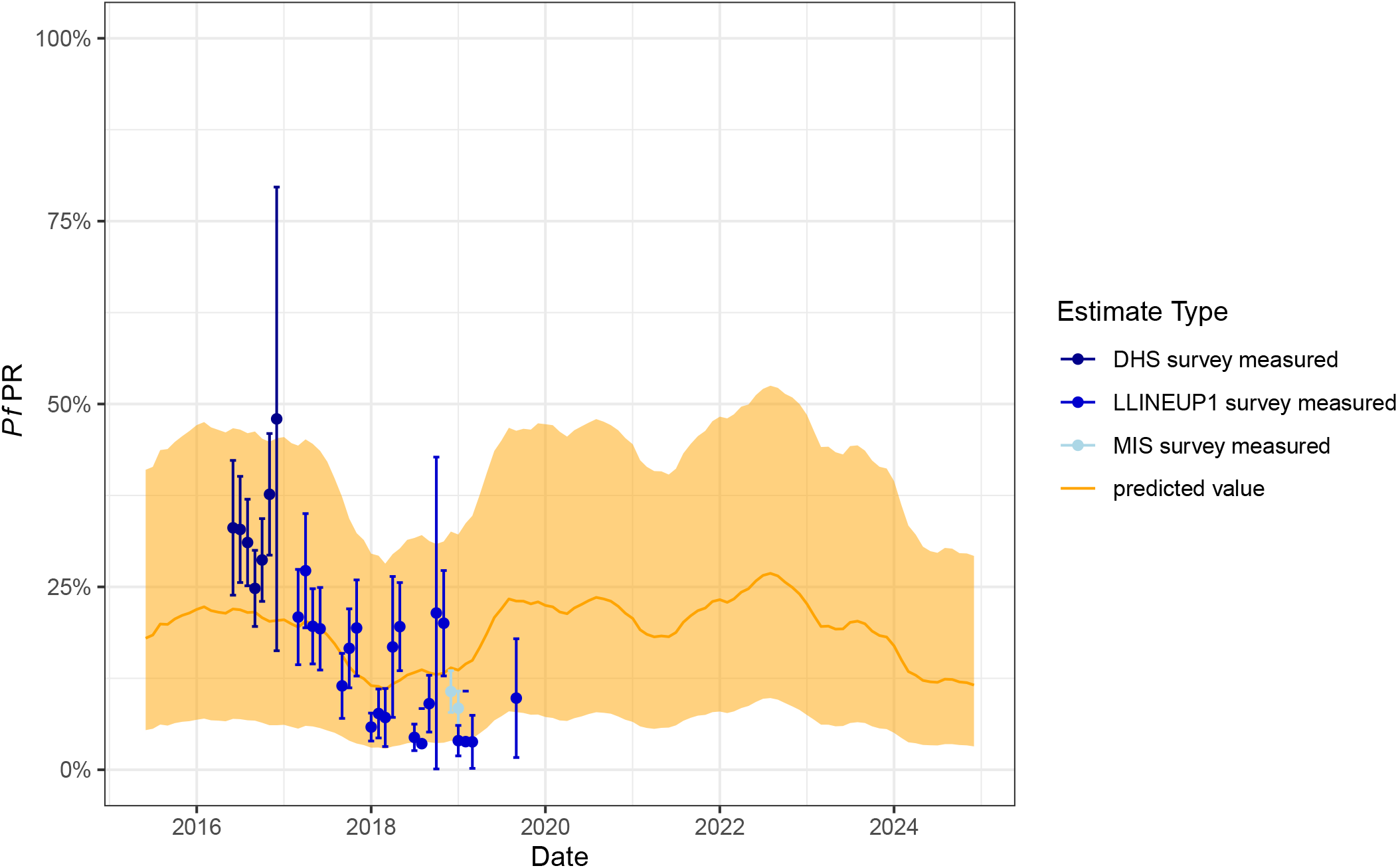
Temporal changes in monthly predicted *Pf*PR from January 2016 to December 2024 for four districts in Uganda. Survey estimates and 95% CI are shown as black dots. Pader, Lira, and Yumbe Districts are all in regions with historically high prevalence, while Kanungu District is in a region with consistently low prevalence.

To visualize monthly variations in the predicted *Pf*PR in Uganda by district, we took a snapshot of one year (2024) and presented the monthly district level changes in predicted *Pf*PR (**Figure 3**). In some regions, like those in the southwest, prevalence was consistently low throughout the year and did not vary greatly between districts. In other regions, such as Karamoja in the Northwest corner, temporal and spatial heterogeneity are observed. This heterogeneity is not apparent when relying on yearly regional estimates of prevalence (**Figure 4**).

**Figure 3.**
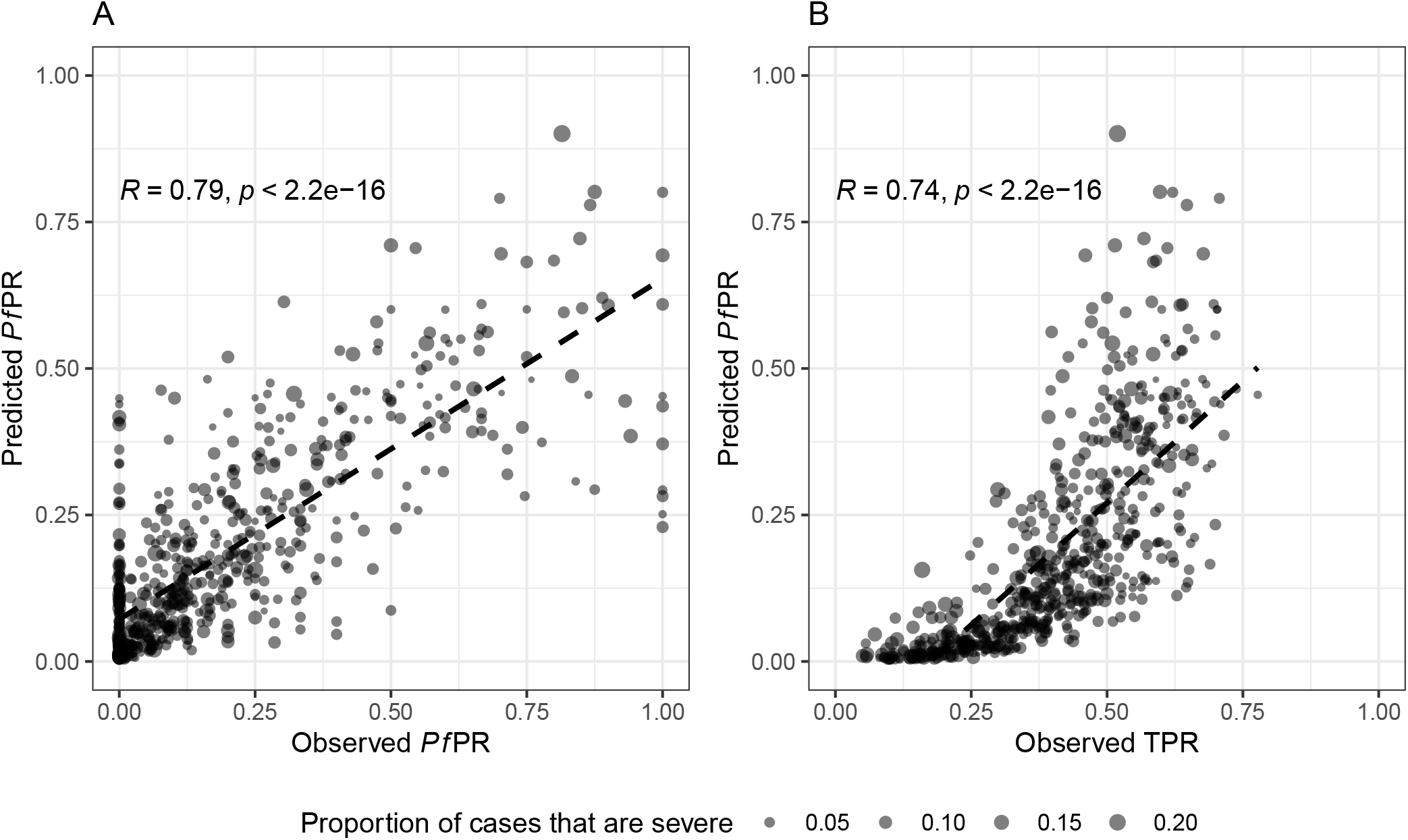
District level monthly estimated *Pf*PR in Uganda for 2024 using HMIS TPR estimates.

**Figure 4.**
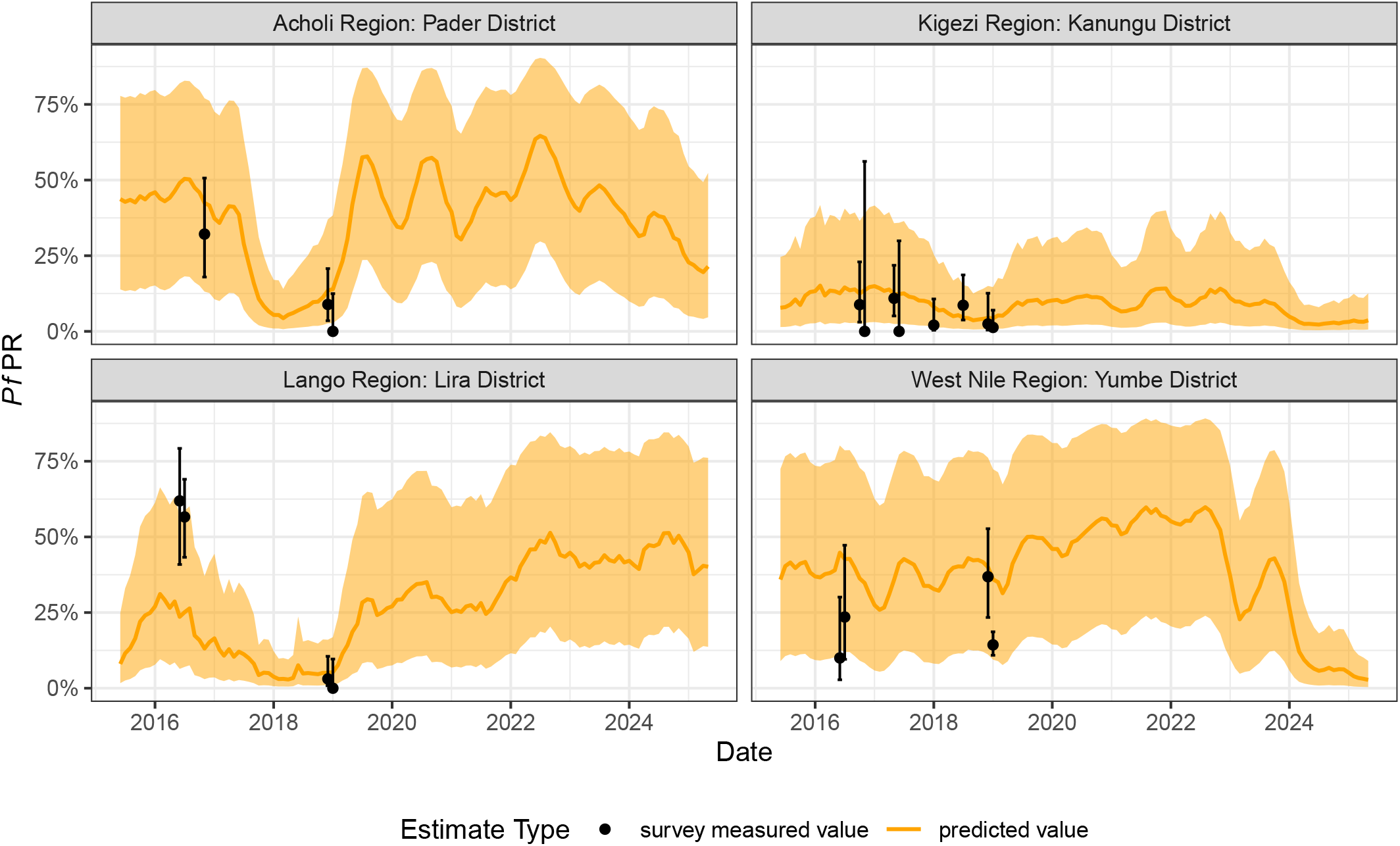
Regional yearly estimates of *Pf*PR in Uganda from 2015-2024 using information from the HMIS.

As another example, Lango region, in the center, has modest prevalence in 2024 at the regional level (26.4%, 95% CI: 8.3%, 55.6%), but district level estimates throughout the year range from a low of 9.2% (95% CI: 7.1%, 11.5%) to a high of 51% (95%CI 20%, 84.5%).

Reviewing the yearly regional estimates highlights a few trends. Prevalence estimates are higher in the northern region, which is known to have higher transmission than the southeast, which consistently has low predicted prevalence. In 2018, a decrease in prevalence in all regions were observed, corresponding with a year of unexplained low prevalence in the country ^10,39^. While the predicted estimates have a large amount of uncertainty, large shifts in prevalence are still observable. West Nile, in the northwest corner of the country, consistently had prevalence estimates >30% until 2024, when the estimate fell to 8.7% (95%CI: 2.9%, 21.6%), corresponding to an intensive vector control strategy in the region.

National monthly trends fall within the survey measured estimates (**Figure 5**), and demonstrate similar seasonality patterns as seen at the district level, as well as the pronounced decrease in prevalence in 2018. Estimates of national level *Pf*PR restricted to the same months and districts as survey data showed similar results. Notably, while the DHS survey estimate fell within the confidence band of our model predicted estimate for 2016, our modeled estimate was about half of the survey estimate.

**Figure 5.**
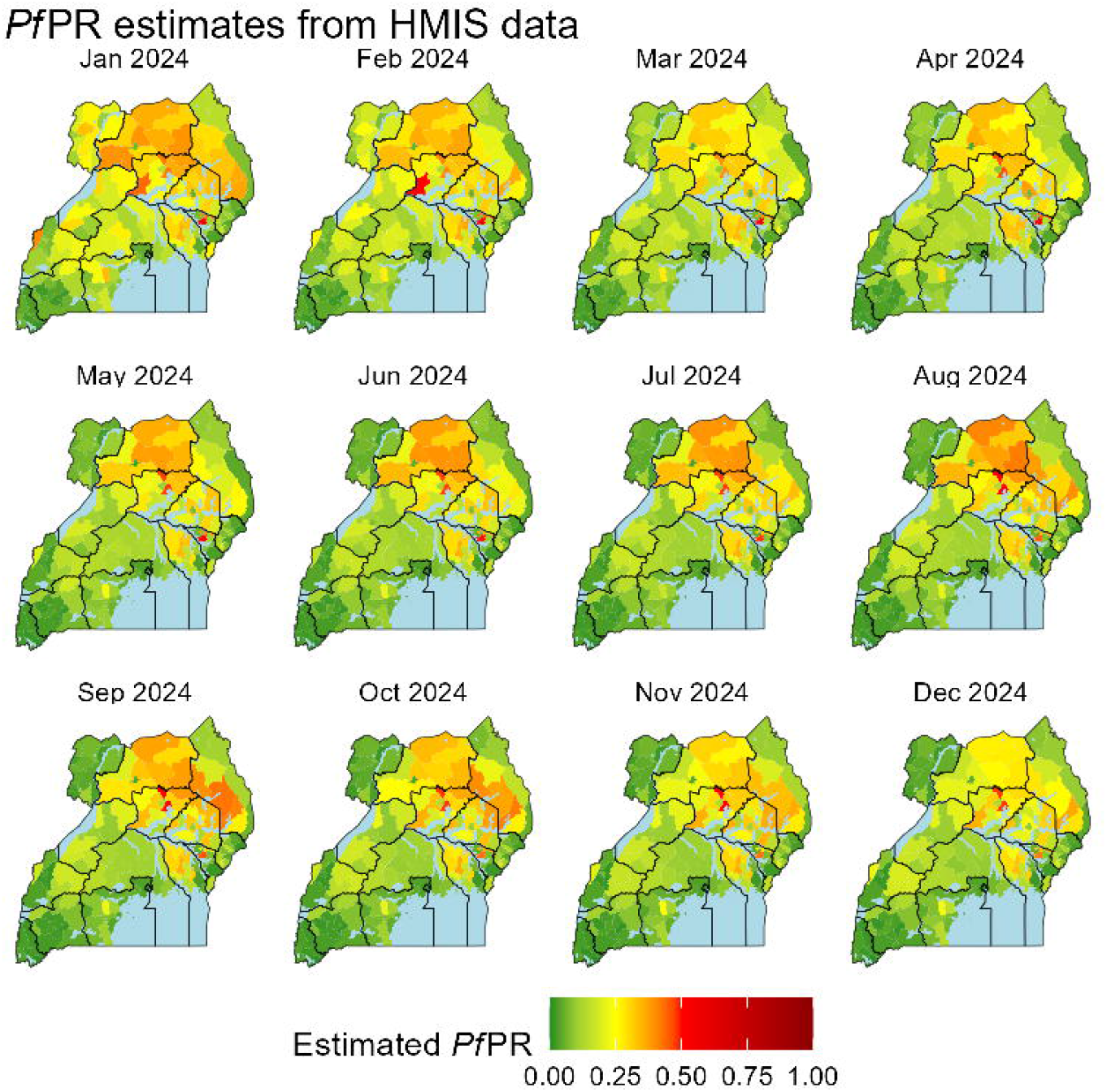
National estimated *Pf*PR by month from model fits (orange line and ribbon) compared with survey estimates for the same months (blue dots with 95% CI).

## Discussion

This study demonstrates the potential of routinely collected health management information system (HMIS) data to generate high-resolution, temporally dynamic estimates of malaria prevalence (*Pf*PR) across Uganda. By calibrating a predictive model using individual-level data from three nationally representative surveys, MIS 2018, DHS 2016, and LLINEUP 2017–2019, we show that test positivity rate (TPR) and the proportion of severe cases from HMIS can reliably estimate *Pf*PR at district, regional, and national levels.

Moderate correlation was observed between TPR and survey-based *Pf*PR, consistent with findings from similar studies in Kenya ^18^. Our final model, which included a 180-day smoothed TPR and proportion of severe cases, achieved strong correlation with observed *Pf*PR (R = 0.79), performing comparably to a more complex model that incorporated environmental covariates (R = 0.81). Given the logistical and financial constraints of integrating external environmental data into routine surveillance, the minimal performance difference suggests that TPR alone captures much of the seasonal variation in transmission. This enhances the feasibility of real-time, HMIS-based monitoring for malaria control and decision-making.

While the model performed well across all three survey datasets, some discrepancies were observed, most notably with the DHS 2016. Although predicted *Pf*PR fell within the confidence bounds of the DHS estimates, the observed *Pf*PR from DHS was nearly twice as high as predicted. It is possible that the discrepancy is due to poorer data quality in HMIS during the DHS survey. In 2016, the HMIS system was in its infancy, and there was continual onboarding and training of health facilities, which could impact data quality. Another possibility is that the difference may be due to the use of rapid diagnostic tests (RDTs) in the DHS, which can overestimate prevalence in high transmission settings by detecting persistent antigens post-clearance ^40,41^. In contrast, MIS and LLINEUP used microscopy, which only detect active infections. Our model, trained on HMIS data incorporating both RDT and microscopy results, may smooth out diagnostic-specific biases but also introduce variability when compared to surveys using a single method. Additionally, DHS had the smallest sample size, with a median of only 8 children tested per cluster and 37 per district, limiting its ability to capture intra-district heterogeneity. MIS and LLINEUP had larger sample sizes and broader geographic coverage, contributing to more stable estimates. These discrepancies highlight the importance of considering diagnostic methods and sampling design when interpreting survey-based malaria metrics, and underscore the value of HMIS-based modeling for generating consistent, scalable estimates.

TPR, while widely used in surveillance, is known to have several limitations that can introduce bias when used to estimate parasite prevalence (*Pf*PR). TPR reflects only individuals who seek care and are tested, missing asymptomatic infections that are common in high-transmission settings ^19^. TPR is also influenced by care-seeking behavior, co-circulating febrile illnesses, diagnostic practices, and intervention coverage ^14,18,42,43^. To mitigate these biases, we implemented several strategies in our modeling approach. We applied temporal smoothing to the TPR using a 180-day rolling average, which helps reduce noise from short-term fluctuations in testing rates and care-seeking patterns ^44,45^. We restricted analysis to facilities with high data completeness, assuming more stable catchment populations and consistent reporting. We also included the proportion of severe cases as a proxy for care-seeking intensity and the presence of other pathogens. Despite these efforts, our model does not account for RDT stockouts, diagnostic algorithm changes, or other febrile illnesses such as dengue or typhoid. Future models could be improved by incorporating data on non-malarial illness prevalence, diagnostic supply chains, and facility catchment populations.

Key strengths of our modeling approach are its ability to uncover spatial and temporal heterogeneity that is obscured in national or regional summaries and to do so in a timely way. District-level predictions revealed seasonal peaks aligned with rainfall patterns and a pronounced dip in prevalence in 2018, consistent with previously reported anomalies. The sharp decline in predicted *Pf*PR in West Nile in 2024, coinciding with intensified vector control efforts, further illustrates the model’s utility for evaluating intervention impact in real-time. Crucially, where model outputs diverge from expected transmission patterns, these can be used to prioritize targets for adaptive malaria control. In these priority areas, the strategy can shift to an iterative intervention process in which targeted data collection and additional studies are deployed to refine predictions, fill knowledge gaps, and allow for a more responsive control effort.

This study has several limitations that should be considered when interpreting the findings. First, the model was trained using survey data restricted to children under five years of age, while HMIS data includes individuals of all ages. The exclusion of older age groups in *Pf*PR estimates may limit understanding of the wider transmission dynamics in an area, particularly given reports of rapid shift in malaria burden to older children and adolescents ^8,46,47^. However, because both model training and prediction were matched using the same age-restricted survey data, we believe this limitation is unlikely to introduce significant inferential bias in the estimates of under 5 *Pf*PR. Finally, the DHS estimates may be farthest from predicted *Pf*PR because it was conducted earlier than the other two surveys, during a period of HMIS onboarding and training, which may have impacted the data quality during this time.

Second, the DHS and MIS surveys were not designed to produce reliable district-level estimates. Sample sizes at the district level were small and limited to a single point in time, which increases uncertainty and limits the ability to capture intra-district heterogeneity. This could affect the precision of model calibration. To mitigate this, we combined data from multiple surveys, which improved sample size and geographic coverage, thereby enhancing the robustness of district-level estimates. Additional adaptive surveys could be developed in the future to improve the accuracy of these models.

Despite these limitations, our modelling approach offers a critical advantage in the current context, where political instability and resource constraints threaten the continuity of large-scale household surveys. As survey data become increasingly infrequent and potentially compromised, reliance on HMIS data for malaria surveillance becomes not only practical but essential. In doing so, we are actively seeking ways of improving the *predictive accuracy* of the models through active and adaptive research. An anticipated side effect of using HMIS facility data to manage malaria is to conduct studies to identify facilities that generate unreliable data, and to identify the underlying causes of those inaccuracies and develop appropriate responses. Our findings support the integration of HMIS-based modeling into national malaria control programs, enabling timely, localized, and cost-effective monitoring of malaria burden.

## Conclusions

This study highlights the feasibility and value of using HMIS data to predict malaria prevalence across Uganda. Our model, calibrated with survey data and validated against observed *Pf*PR, provides accurate, high-resolution estimates that capture spatial and temporal heterogeneity in transmission. In the face of declining survey frequency and growing political uncertainty, HMIS-based modeling offers a resilient and cost-effective alternative for malaria surveillance. Strengthening routine data systems and integrating predictive analytics into national malaria strategies will be critical to sustaining progress toward malaria control and elimination.

## Data Availability

All data produced in the present study are available upon reasonable request to the authors.

## Abbreviations

AIC: Akaike Information Criterion
DHS: demographic health survey
HMIS: Health Management Information System
IRS: Indoor residual spraying
ITN: Insecticide treated bed-net
LLIN: long-lasting insecticidal net
LLINEUP: LLIN Evaluation in Uganda Project
MIS: Malaria Indicator Survey
MOH: Ministry of Health
NMCD: National Malaria Control Division
*Pf*PR: *Plasmodium falciparum* Parasite Rate
TPR: Test Positivity Rate
UMRESP: Uganda Malaria Reduction Strategic Plan
WHO: World Health Organization

## Declarations

Ethical approval for study procedures and data collection was not required due to the due to the de-identified nature of the data.

## Authors’ contributions

Conceptualization: JO, JR,ARC,DLS,DEBH; Funding acquisition: DLS; Methodology: JO, JR, ARC, DLS,DH; Investigation: JO, JR, ARC,DLS, DEBH; Data curation: JO, JR, ARC,TE,DLS, DEBH; Formal analysis: JO, JR,ARC,DLS, DEBH; Writing – original draft: JO, DEBH; Writing – review & editing: JO, JR, ARC,DM,JNN,MR,CMS,PM,DLS, DEBH; All authors read and approved the final manuscript.

## Acknowledgements

We would like to thank the entire study team and the administration of the Pilgrim Africa, Department of Health Information and NMCD at the Ministry of Health for all their contributions.

## References

1. World Malaria Report 2025: Addressing the Threat of Antimalarial Drug Resistance. Geneva: World Health Organization; 2025. Licence: CC BY-NC-SA 3.0 IGO.

2. Subnational Tailoring of Malaria Strategies and Interventions: Reference Manual. Geneva: World Health Organization; 2025. Licence: CC BY-NC-SA 3.0 IGO.

3. Hay SI, Smith DL, Snow RW. Measuring malaria endemicity from intense to interrupted transmission. The Lancet Infectious Diseases. 2008;8(6):369–378. doi:10.1016/S1473-3099(08)70069-0

4. Tusting LS, Bousema T, Smith DL, Drakeley C. Measuring Changes in Plasmodium falciparum Transmission. In: Advances in Parasitology. Vol 84. Elsevier; 2014:151–208. doi:10.1016/B978-0-12-800099-1.00003-X

5. Amoah B, McCann RS, Kabaghe AN, et al. Identifying Plasmodium falciparum transmission patterns through parasite prevalence and entomological inoculation rate. eLife. 2021;10:e65682. doi:10.7554/eLife.65682

6. Guerra CA, Hay SI, Lucioparedes LS, et al. Assembling a global database of malaria parasite prevalence for the Malaria Atlas Project. Malar J. 2007;6(1):17. doi:10.1186/1475-2875-6-17

7. Mappin B, Cameron E, Dalrymple U, et al. Standardizing Plasmodium falciparum infection prevalence measured via microscopy versus rapid diagnostic test. Malar J. 2015;14(1):460. doi:10.1186/s12936-015-0984-9

8. Kigozi SP, Kigozi RN, Epstein A, et al. Rapid shifts in the age-specific burden of malaria following successful control interventions in four regions of Uganda. Malar J. 2020;19(1):1. doi:10.1186/s12936-020-03196-7

9. Hay SI, Snow RW. The Malaria Atlas Project: Developing Global Maps of Malaria Risk. PLoS Med. 2006;3(12):e473. doi:10.1371/journal.pmed.0030473

10. Kigozi SP, Kigozi RN, Sebuguzi CM, et al. Spatial-temporal patterns of malaria incidence in Uganda using HMIS data from 2015 to 2019. BMC Public Health. 2020;20(1):1913. doi:10.1186/s12889-020-10007-w

11. Ssempiira J, Nambuusi B, Kissa J, et al. Geostatistical modelling of malaria indicator survey data to assess the effects of interventions on the geographical distribution of malaria prevalence in children less than 5 years in Uganda. PLOS ONE. 2017;12(4):e0174948. doi:10.1371/journal.pone.0174948

12. Kudymowa, J., Van Schoubroeck, C., Leow, A., & Basnak, M. 2025. Data Systems for Malaria Burden Estimation: Challenges, Initiatives, and Opportunities. Rethink Priorities. Https://Rethinkpriorities.Org/Research-Area/Data-Systems-for-Malaria/.

13. Okiring J, Epstein A, Namuganga JF, et al. Relationships between test positivity rate, total laboratory confirmed cases of malaria, and malaria incidence in high burden settings of Uganda: an ecological analysis. Malar J. 2021;20(1):1. doi:10.1186/s12936-021-03584-7

14. Boyce RM, Reyes R, Matte M, et al. Practical Implications of the Non-Linear Relationship between the Test Positivity Rate and Malaria Incidence. Culleton R, ed. PLOS ONE. 2016;11(3):3. doi:10.1371/journal.pone.0152410

15. Kigozi SP, Kigozi RN, Sserwanga A, et al. Malaria Burden through Routine Reporting: Relationship between Incidence and Test Positivity Rates. The American Journal of Tropical Medicine and Hygiene. 2019;101(1):1. doi:10.4269/ajtmh.18-0901

16. Raouf S, Mpimbaza A, Kigozi R, et al. Resurgence of malaria following discontinuation of indoor residual spraying of insecticide in a previously high transmission intensity area of Uganda. Clin Infect Dis. Published online March 24, 2017. doi:10.1093/cid/cix251

17. Jensen TP, Bukirwa H, Njama-Meya D, et al. Use of the slide positivity rate to estimate changes in malaria incidence in a cohort of Ugandan children. Malar J. 2009;8(1):1. doi:10.1186/1475-2875-8-213

18. Kamau A, Mtanje G, Mataza C, Malla L, Bejon P, Snow RW. The relationship between facility-based malaria test positivity rate and community-based parasite prevalence. Braga ÉM, ed. PLoS ONE. 2020;15(10):e0240058. doi:10.1371/journal.pone.0240058

19. Stresman G, Sepúlveda N, Fornace K, et al. Association between the proportion of Plasmodium falciparum and Plasmodium vivax infections detected by passive surveillance and the magnitude of the asymptomatic reservoir in the community: a pooled analysis of paired health facility and community data. The Lancet Infectious Diseases. 2020;20(8):953–963. doi:10.1016/S1473-3099(20)30059-1

20. Epstein A, Maiteki-Sebuguzi C, Namuganga JF, et al. Resurgence of malaria in Uganda despite sustained indoor residual spraying and repeated long lasting insecticidal net distributions. Ashton R, ed. PLOS Global Public Health. 2022;2(9):9. doi:10.1371/journal.pgph.0000676

21. Kamya MR, Nankabirwa JI, Arinaitwe E, et al. Dramatic resurgence of malaria after 7 years of intensive vector control interventions in Eastern Uganda. Ashton R, ed. PLOS Glob Public Health. 2024;4(8):e0003254. doi:10.1371/journal.pgph.0003254

22. Ocen E, Opito R, Tegu C, Oula A, Olupot-Olupot P. Severe malaria burden, clinical spectrum and outcomes at Apac district hospital, Uganda: a retrospective study of routine health facility-based data. Malar J. 2023;22(1):325. doi:10.1186/s12936-023-04761-6

23. Epstein A, Namuganga JF, Kamya EV, et al. Estimating malaria incidence from routine health facility-based surveillance data in Uganda. Malaria Journal. 2020;19(1):1. doi:10.1186/s12936-020-03514-z

24. Simple O, Mindra A, Obai G, Ovuga E, Odongo-Aginya EI. Influence of Climatic Factors on Malaria Epidemic in Gulu District, Northern Uganda: A 10-Year Retrospective Study. Malar Res Treat. 2018;2018:5482136. doi:10.1155/2018/5482136

25. Katusiime M, Kabwama SN, Rukundo G, et al. Malaria outbreak facilitated by engagement in activities near swamps following increased rainfall and limited preventive measures: Oyam District, Uganda. Ashton R, ed. PLOS Global Public Health. 2022;2(8):8. doi:10.1371/journal.pgph.0000239

26. Okello PE, Van Bortel W, Byaruhanga AM, et al. Variation in malaria transmission intensity in seven sites throughout Uganda. Am J Trop Med Hyg. 2006;75(2):2.

27. Kamya MR, Arinaitwe E, Wanzira H, et al. Malaria Transmission, Infection, and Disease at Three Sites with Varied Transmission Intensity in Uganda: Implications for Malaria Control. The American Society of Tropical Medicine and Hygiene. 2015;92(5):903–912. doi:10.4269/ajtmh.14-0312

28. Nuwa A, Baker K, Bonnington C, et al. A non-randomized controlled trial to assess the protective effect of SMC in the context of high parasite resistance in Uganda. Malar J. 2023;22(1):63. doi:10.1186/s12936-023-04488-4

29. Kwiringira A, Kwesiga B, Migisha R, et al. Effect of seasonal malaria chemoprevention on incidence of malaria among children under five years in Kotido and Moroto Districts, Uganda, 2021: time series analysis. Malar J. 2024;23(1):389. doi:10.1186/s12936-024-05220-6

30. Uganda Bureau of Statistics (UBOS) and ICF. 2018. Uganda Demographic and Health Survey 2016. Kampala, Uganda and Rockville, Maryland, USA: UBOS and ICF.

31. Uganda National Malaria Control Division (NMCD), Uganda Bureau of Statistics (UBOS), and ICF. 2020. Uganda Malaria Indicator Survey 2018-19. Kampala, Uganda, and Rockville, Maryland, USA: NMCD, UBOS, and ICF. 190.

32. Gonahasa S, Maiteki-Sebuguzi C, Rugnao S, et al. LLIN Evaluation in Uganda Project (LLINEUP): factors associated with ownership and use of long-lasting insecticidal nets in Uganda: a cross-sectional survey of 48 districts. Malar J. 2018;17(1):1. doi:10.1186/s12936-018-2571-3

33. Staedke SG, Gonahasa S, Dorsey G, et al. Effect of long-lasting insecticidal nets with and without piperonyl butoxide on malaria indicators in Uganda (LLINEUP): a pragmatic, cluster-randomised trial embedded in a national LLIN distribution campaign. The Lancet. 2020;395(10232):10232. doi:10.1016/S0140-6736(20)30214-2

34. Staedke SG, Kamya MR, Dorsey G, et al. LLIN Evaluation in Uganda Project (LLINEUP) – Impact of long-lasting insecticidal nets with, and without, piperonyl butoxide on malaria indicators in Uganda: study protocol for a cluster-randomised trial. Trials. 2019;20(1):1. doi:10.1186/s13063-019-3382-8

35. Funk C, Peterson P, Landsfeld M, et al. The climate hazards infrared precipitation with stations— a new environmental record for monitoring extremes. Sci Data. 2015;2(1):1. doi:10.1038/sdata.2015.66

36. National Aeronautics and Space Administration (NASA). Moderate Resolution Imaging Spectroradiometer. https://modis.gsfc.nasa.gov/data/

37. Uganda Bureau of Statistics 2024: The National Population and Housing Census 2024 – Final Report - Volume 1 (Main), Kampala, Uganda.

38. Weiss DJ, Nelson A, Gibson HS, et al. A global map of travel time to cities to assess inequalities in accessibility in 2015. Nature. 2018;553(7688):333–336. doi:10.1038/nature25181

39. Oumo PO, Thiwe P, Kadobera D, Ario AR. Trends and distribution of malaria deaths among the general population, Uganda 2015-2019. https://uniph.go.ug/trends-and-distribution-of-malaria-deaths-among-the-general-population-uganda-2015-2019/

40. Dalrymple U, Arambepola R, Gething PW, Cameron E. How long do rapid diagnostic tests remain positive after anti-malarial treatment? Malar J. 2018;17(1):228. doi:10.1186/s12936-018-2371-9

41. Sheahan W, Golden A, Barney R, et al. Estimating malaria antigen dynamics and the time to negativity of next-generation malaria rapid diagnostic tests. Malar J. 2025;24(1):109. doi:10.1186/s12936-025-05350-5

42. Sinzinkayo D, Baza D, Gnanguenon V, Koepfli C. The lead-up to epidemic transmission: malaria trends and control interventions in Burundi 2000 to 2019. Malar J. 2021;20(1):298. doi:10.1186/s12936-021-03830-y

43. Bisanzio D, Keita MS, Camara A, et al. Malaria trends in districts that were targeted and not-targeted for seasonal malaria chemoprevention in children under 5 years of age in Guinea, 2014–2021. BMJ Glob Health. 2024;9(2):e013898. doi:10.1136/bmjgh-2023-013898

44. Hussien HH, Eissa FH, Awadalla KE. Statistical Methods for Predicting Malaria Incidences Using Data from Sudan. Malaria Research and Treatment. 2017;2017:1–9. doi:10.1155/2017/4205957

45. Dieng S, Adebayo-Ojo TC, Kruger T, et al. Geo-epidemiology of malaria incidence in the Vhembe District to guide targeted elimination strategies, South-Africa, 2015–2018: a local resurgence. Sci Rep. 2023;13(1):11049. doi:10.1038/s41598-023-38147-0

46. Pemberton-Ross P, Smith TA, Hodel EM, Kay K, Penny MA. Age-shifting in malaria incidence as a result of induced immunological deficit: a simulation study. Malar J. 2015;14(1):287. doi:10.1186/s12936-015-0805-1

47. Griffin JT, Ferguson NM, Ghani AC. Estimates of the changing age-burden of Plasmodium falciparum malaria disease in sub-Saharan Africa. Nature Communications. 2014;5. doi:10.1038/ncomms4136

