## Supplemental Table 1 for "Estimating *Plasmodium falciparum* Parasite Rate using Test Positivity Rate from 2016-2024: Health Management Information Systems in Uganda"

**Supplemental Table 1.** Akaike Information Criterion (AIC) for models assessing the relationship between a 180-day smoothed test positive rate (TPR) and *Pf*PR with various covariates added in. All models were generalized linear mixed models with a logit link function with the following form (r=region, d=district, t= time in months):

$$logit\left( {pos\_pf}_{idrt} \right)= \beta_{0}+\beta_{1}{TPR}_{irdt}+\sum_{k}^{n} \beta_{k}Z_{kidrt}+\alpha_{rd}+\gamma_{t}$$

| **Independent Variable** | **Additional covariates** | **AIC** |
| --- | --- | --- |
| TPR (180-day smooth) | prop_sev + temperature + rain | 19838.9 |
| TPR (180-day smooth) | prop_sev + temperature + rain + pop_density | 19839.6 |
| TPR (180-day smooth) | prop_sev + temperature | 19841.9 |
| TPR (180-day smooth) | rain + temperature | 19899.2 |
| TPR (180-day smooth) | rain + temperature | 19899.2 |
| TPR (180-day smooth) | temperature | 19901.6 |
| TPR (180-day smooth) | temperature + pop_density | 19902.0 |
| TPR (180-day smooth) | prop_sev + rain + pop_density + weighted_median_walk_time | 19910.7 |
| TPR (180-day smooth) | prop_sev + rain + pop_density + weighted_median_walk_time | 19910.7 |
| TPR (180-day smooth) | prop_sev + rain | 19915.2 |
| TPR (180-day smooth) | prop_sev + temp_effect + rain | 19916.5 |
| TPR (180-day smooth) | prop_sev + temp_effect + rain + pop_density | 19917.3 |
| TPR (180-day smooth) | prop_sev + weighted_median_walk_time | 19918.1 |
| TPR (180-day smooth) | prop_sev + temp_effect | 19921.3 |
| TPR (180-day smooth) | prop_sev | 19924.0 |
| TPR (180-day smooth) | weighted_median_walk_time + rain | 19975.4 |
| TPR (180-day smooth) | rain | 19978.8 |
| TPR (180-day smooth) | rain + temp_effect | 19979.7 |
| TPR (180-day smooth) | prop_lt_20_wtd | 19984.9 |
| TPR (180-day smooth) | temp_effect | 19985.7 |
| TPR (180-day smooth) | temp_effect + pop_density | 19986.5 |
| TPR (180-day smooth) | weighted_median_walk_time | 19987.0 |
| TPR (180-day smooth) |  | 19989.9 |
| TPR (180-day smooth) | pop_density | 19990.7 |

prop_sev = proportion of all cases in the district that were severe

temperature =average monthly land surface temperature in district

rain = average monthly rainfall in district

pop_density = population density in district

weight_median_walk_time = population weight median walk time to nearest health facility. Walk time calculated for 1kmx1km grids within district.

temp_effect = Gaussian transformation of raw temperature data, assuming thermal optimum for transmission centered at 25^o^C with symmetric decline in suitability as temperatures deviate.
