## Supplemental Figure 1 for "Estimating *Plasmodium falciparum* Parasite Rate using Test Positivity Rate from 2016-2024: Health Management Information Systems in Uganda"

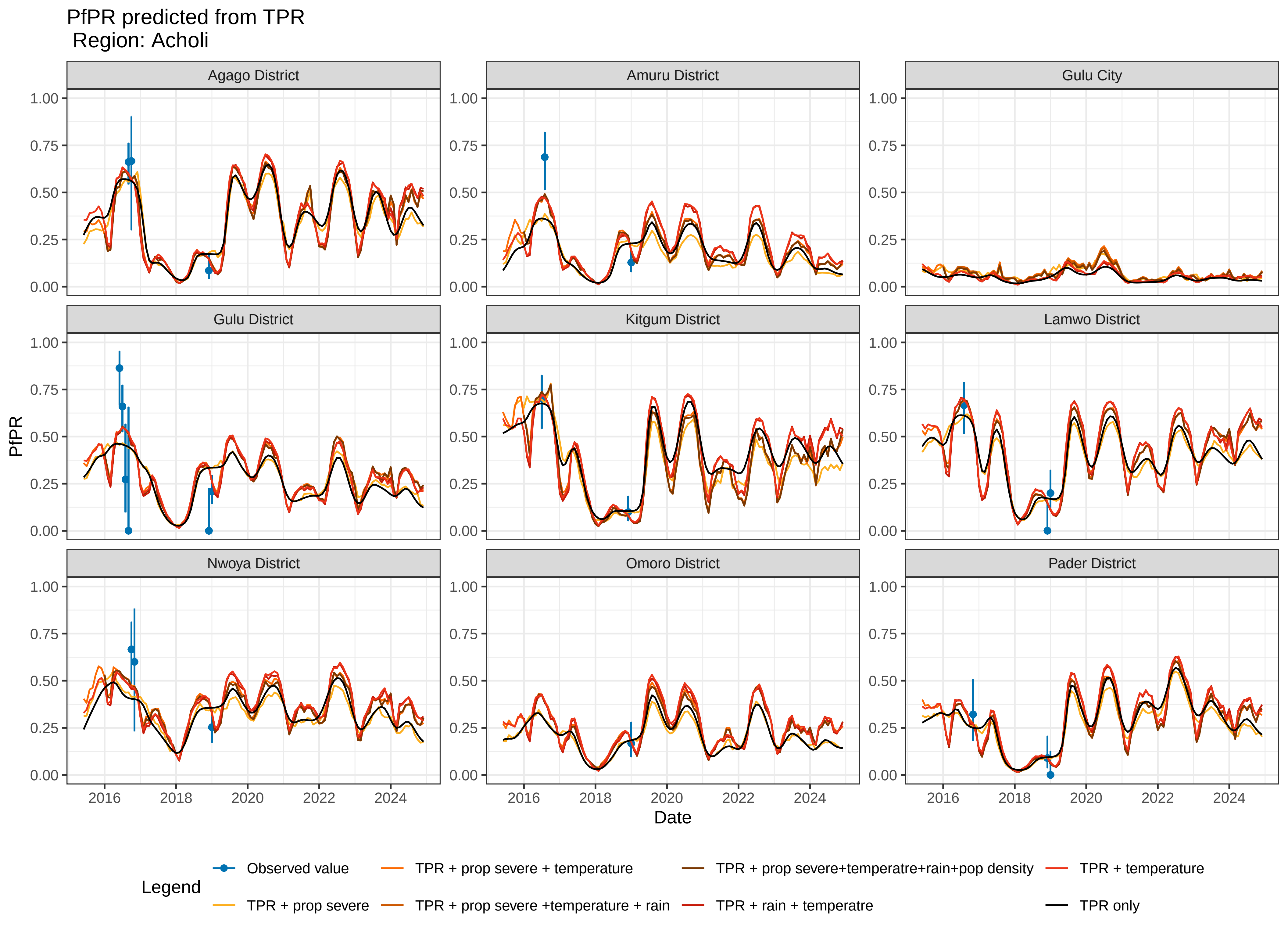

PfPR predicted from TPR
Region: Acholi
Agago District
Amuru District
Gulu City
Gulu District
Kitgum District
Lamwo District
Nwoya District
Omoro District
Pader District
PfPR
1.00
0.75
0.50
0.25
0.00
2016
2018
2020
2022
2024
Date
Legend
Observed value
TPR + prop severe
TPR + prop severe + temperature
TPR + prop severe +temperature + rain
TPR + prop severe+temperatre+rain+pop density
TPR + rain + temperatre
TPR + temperature
TPR only

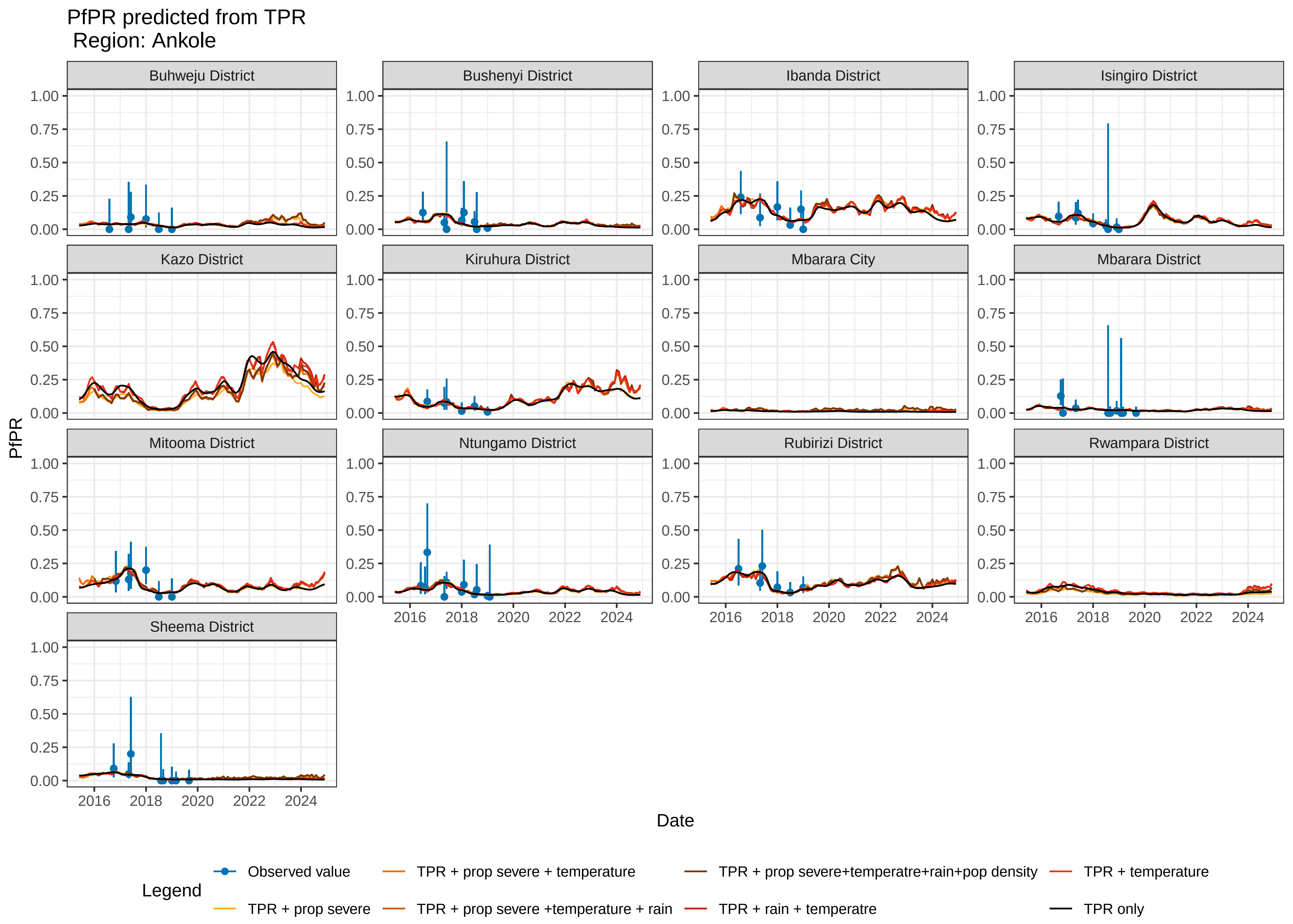
PfPR predicted from TPR
Region: Ankole
Buhweju District
Bushenyi District
Ibanda District
Isingiro District
Kazo District
Kiruhura District
Mbarara City
Mbarara District
Mitooma District
Ntungamo District
Rubirizi District
Rwampara District
Sheema District
1.00
0.75
0.50
0.25
0.00
2016
2018
2020
2022
2024
PfPR
Date
Legend
Observed value
TPR + prop severe
TPR + prop severe + temperature
TPR + prop severe +temperature + rain
TPR + prop severe+temperatre+rain+pop density
TPR + rain + temperatre
TPR + temperature
TPR only

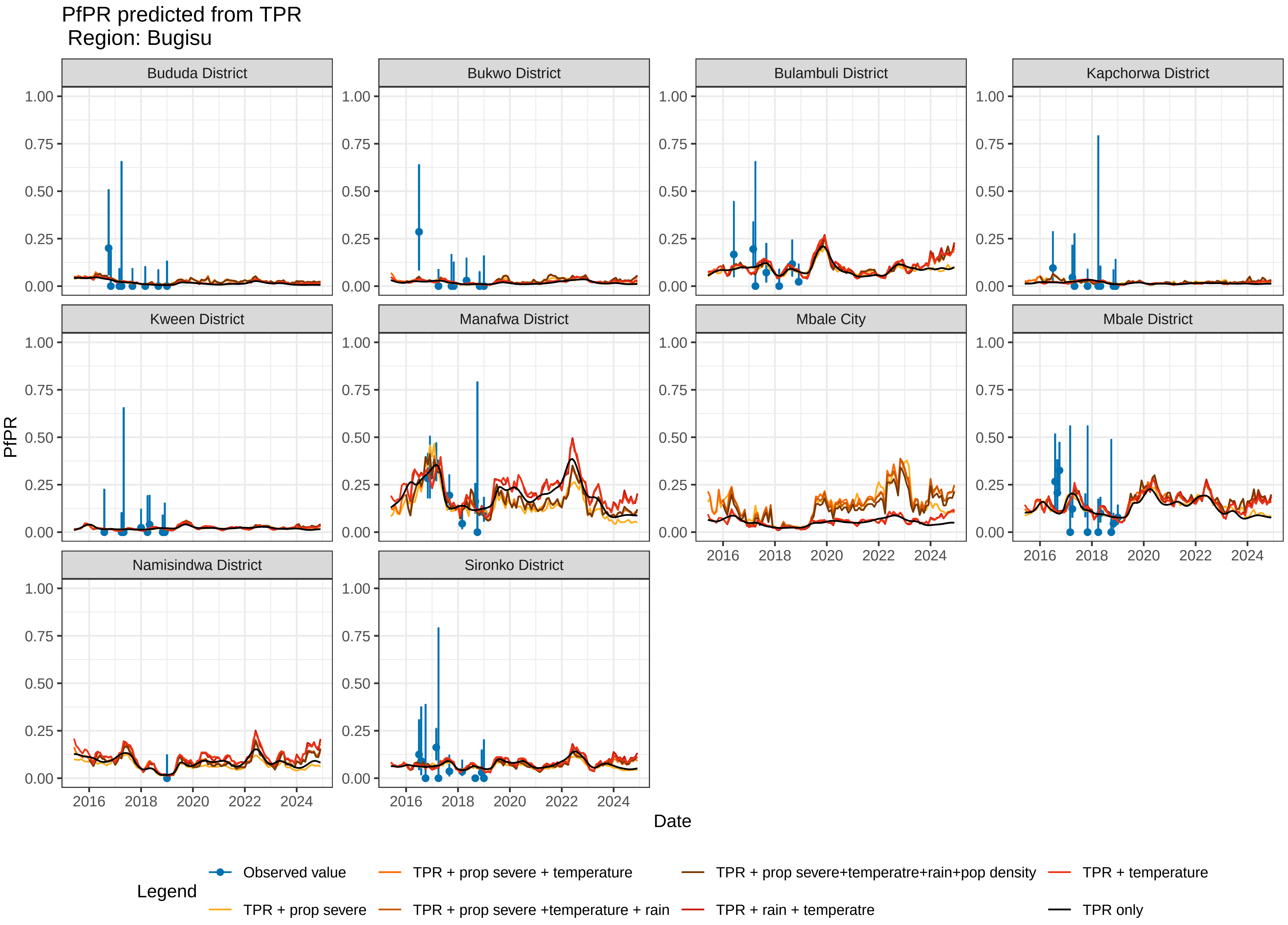
PfPR predicted from TPR
Region: Bugisu
Bududa District
Bukwo District
Bulambuli District
Kapchorwa District
Kween District
Manafwa District
Mbale City
Mbale District
Namisindwa District
Sironko District
PfPR
1.00
0.75
0.50
0.25
0.00
2016
2018
2020
2022
2024
Date
Legend
Observed value
TPR + prop severe
TPR + prop severe + temperature
TPR + prop severe +temperature + rain
TPR + prop severe+temperatre+rain+pop density
TPR + rain + temperatre
TPR + temperature
TPR only

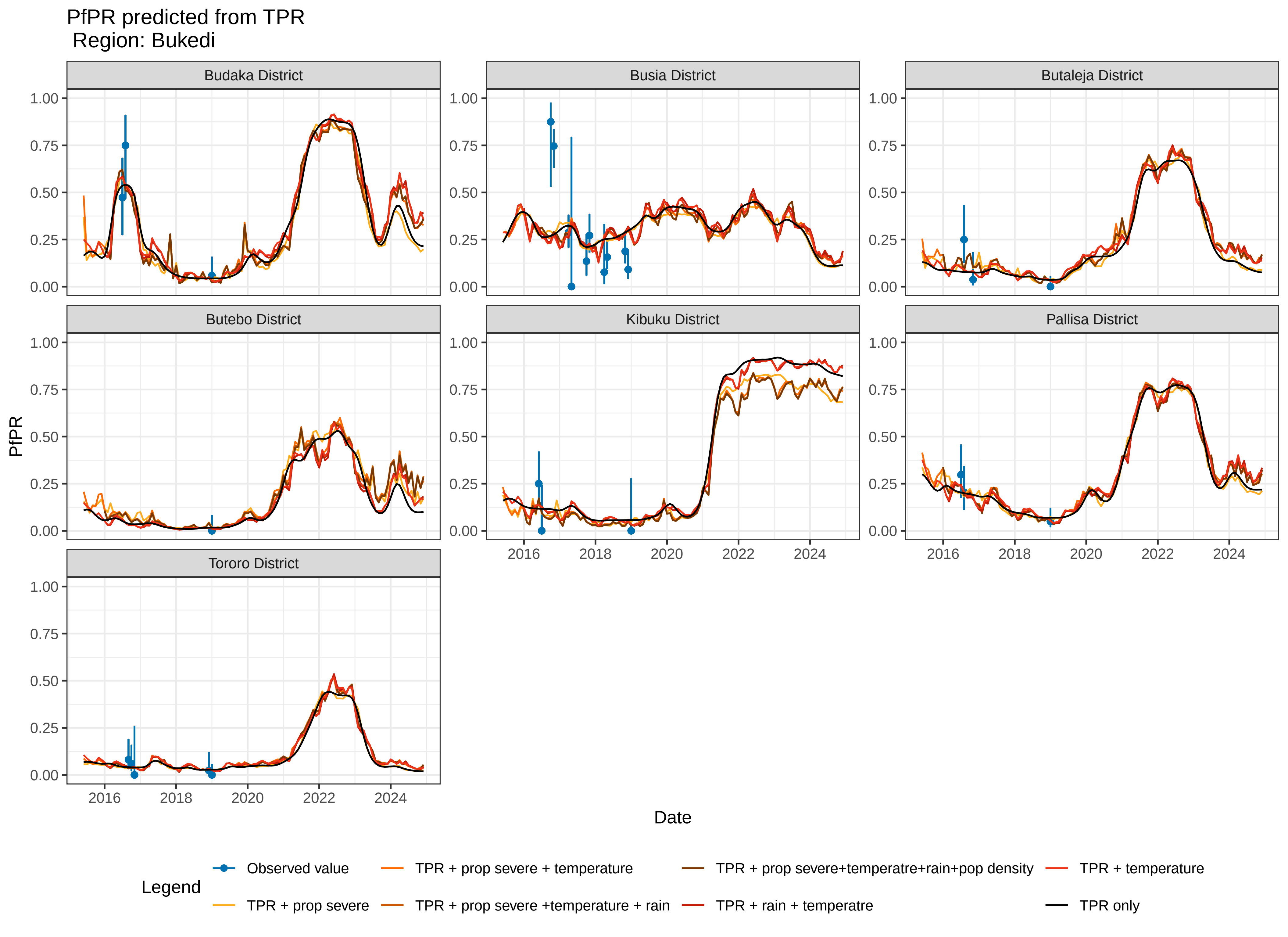
PfPR predicted from TPR
Region: Bukedi
Budaka District
Busia District
Butaleja District
Butebo District
Kibuku District
Pallisa District
Tororo District
PfPR
Date
1.00
0.75
0.50
0.25
0.00
2016
2018
2020
2022
2024
Legend
Observed value
TPR + prop severe
TPR + prop severe + temperature
TPR + prop severe +temperature + rain
TPR + prop severe+temperatre+rain+pop density
TPR + rain + temperatre
TPR + temperature
TPR only

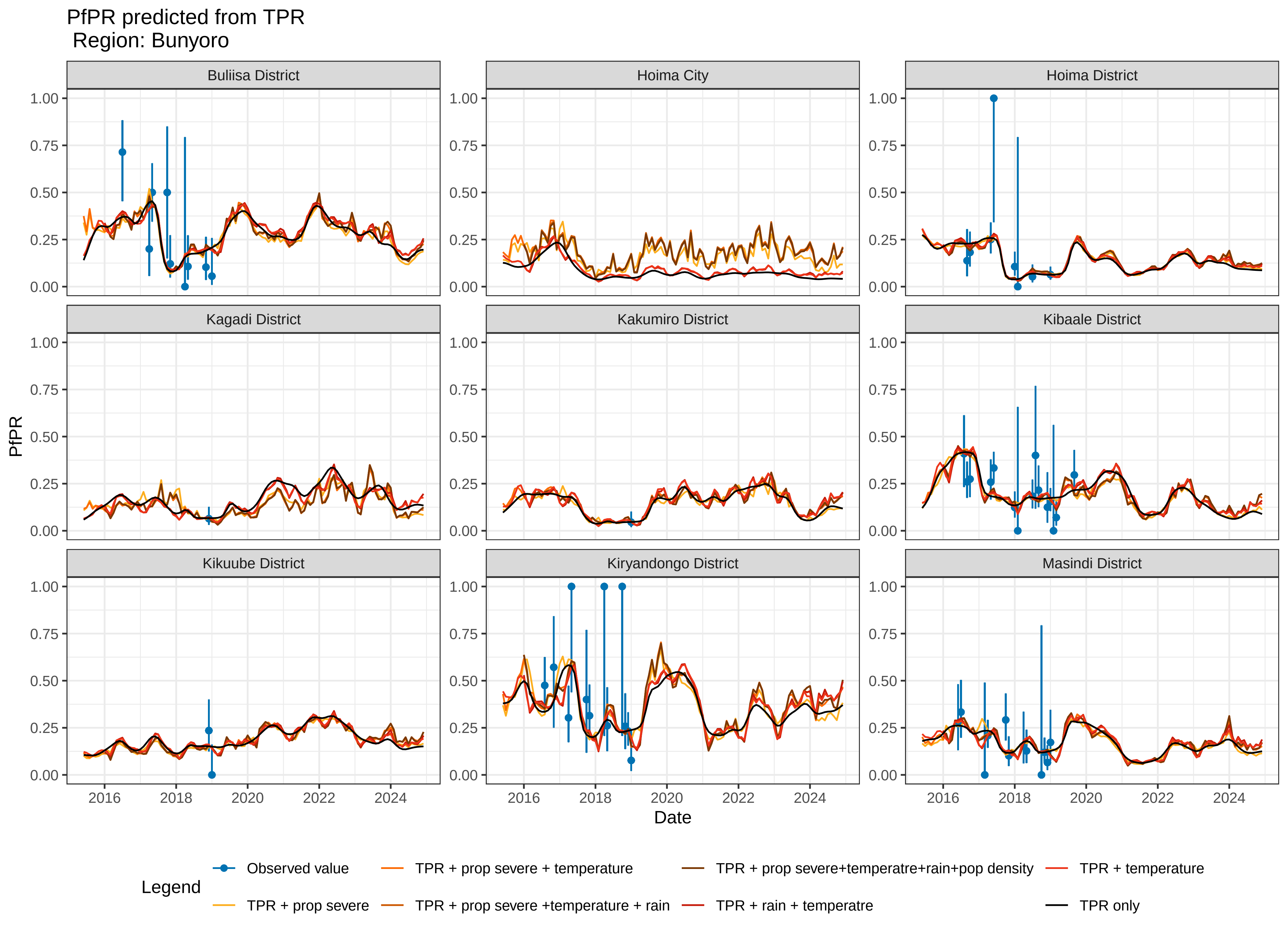

PfPR predicted from TPR
Region: Bunyoro
Buliisa District
Hoima City
Hoima District
Kagadi District
Kakumiro District
Kibaale District
Kikuube District
Kiryandongo District
Masindi District
PfPR
Date
0.00
0.25
0.50
0.75
1.00
2016
2018
2020
2022
2024
Legend
Observed value
TPR + prop severe
TPR + prop severe + temperature
TPR + prop severe +temperature + rain
TPR + prop severe+temperatre+rain+pop density
TPR + rain + temperatre
TPR + temperature
TPR only

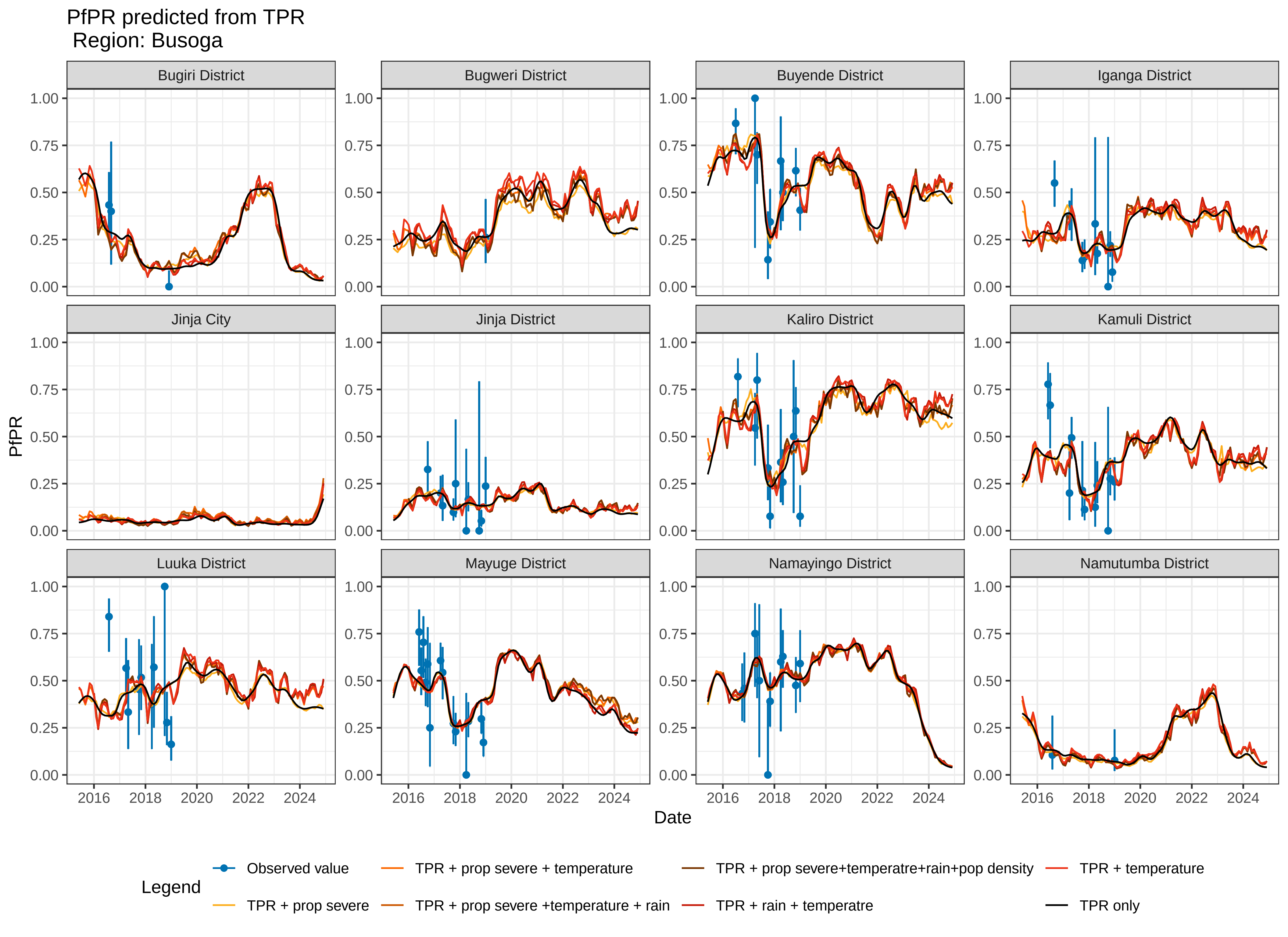

PfPR predicted from TPR
Region: Busoga
Bugiri District
Bugweri District
Buyende District
Iganga District
Jinja City
Jinja District
Kaliro District
Kamuli District
Luuka District
Mayuge District
Namayingo District
Namutumba District
PfPR
1.00
0.75
0.50
0.25
0.00
Date
2016
2018
2020
2022
2024
Legend
Observed value
TPR + prop severe
TPR + prop severe + temperature
TPR + prop severe +temperature + rain
TPR + prop severe+temperatre+rain+pop density
TPR + rain + temperatre
TPR + temperature
TPR only

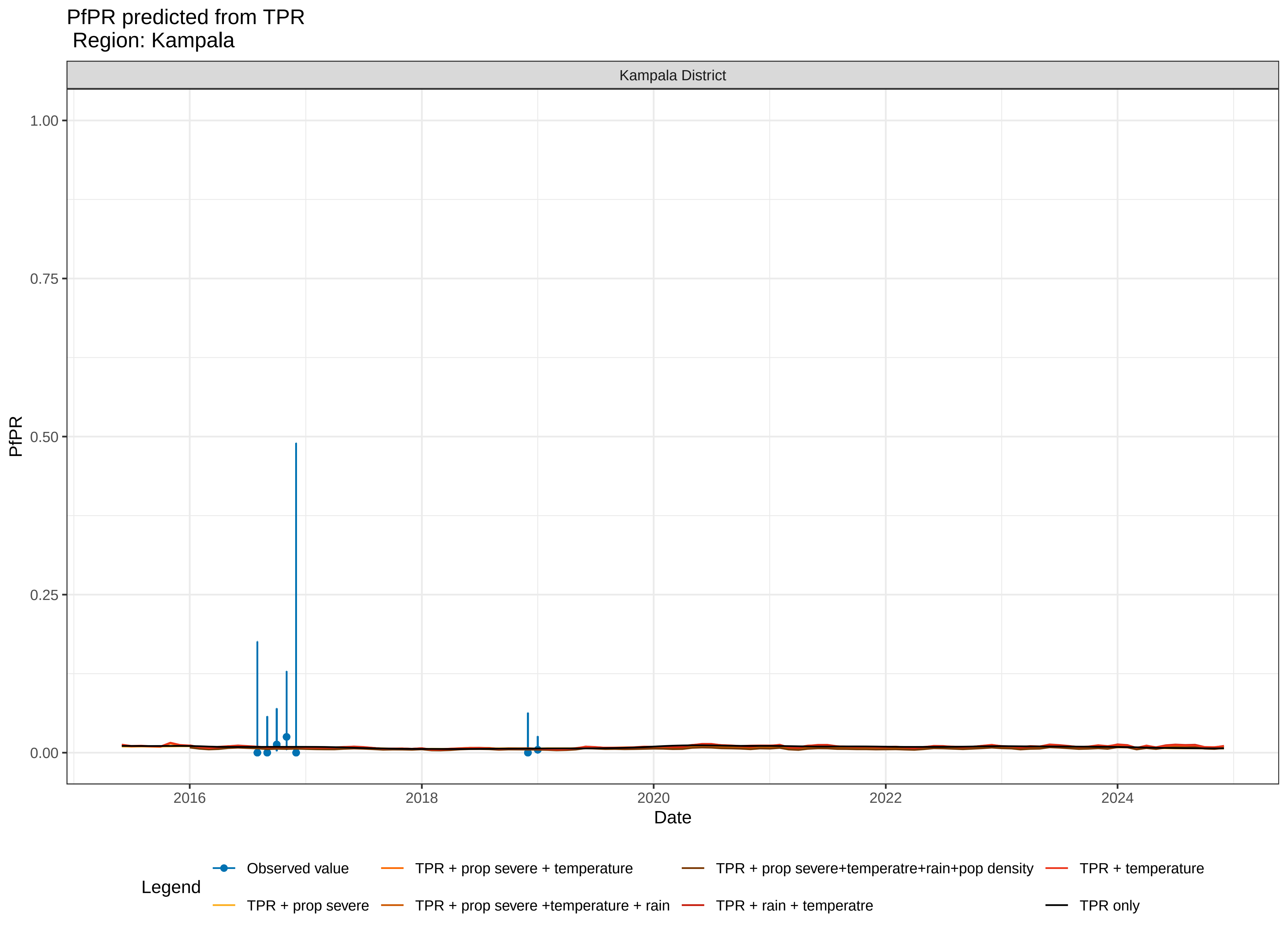

PfPR predicted from TPR
Region: Kampala
Kampala District
1.00
0.75
0.50
0.25
0.00
PfPR
2016
2018
2020
2022
2024
Date
Legend
Observed value
TPR + prop severe
TPR + prop severe + temperature
TPR + prop severe +temperature + rain
TPR + prop severe+temperatre+rain+pop density
TPR + rain + temperatre
TPR + temperature
TPR only

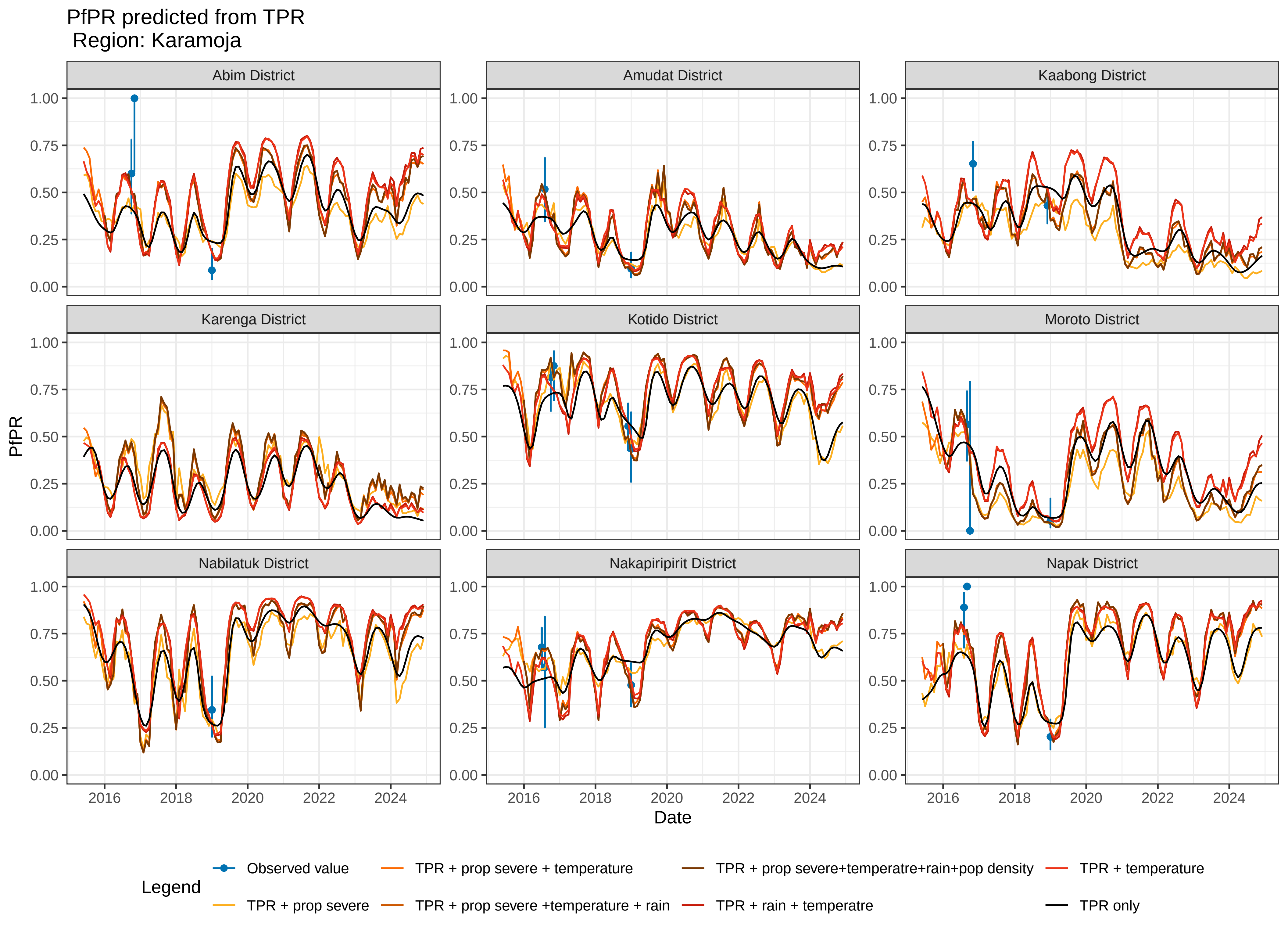

PfPR predicted from TPR
Region: Karamoja
Abim District
Amudat District
Kaabong District
Karenga District
Kotido District
Moroto District
Nabilatuk District
Nakapiripirit District
Napak District
PfPR
Date
1.00
0.75
0.50
0.25
0.00
2016
2018
2020
2022
2024
Legend
Observed value
TPR + prop severe
TPR + prop severe + temperature
TPR + prop severe +temperature + rain
TPR + prop severe+temperatre+rain+pop density
TPR + rain + temperatre
TPR + temperature
TPR only

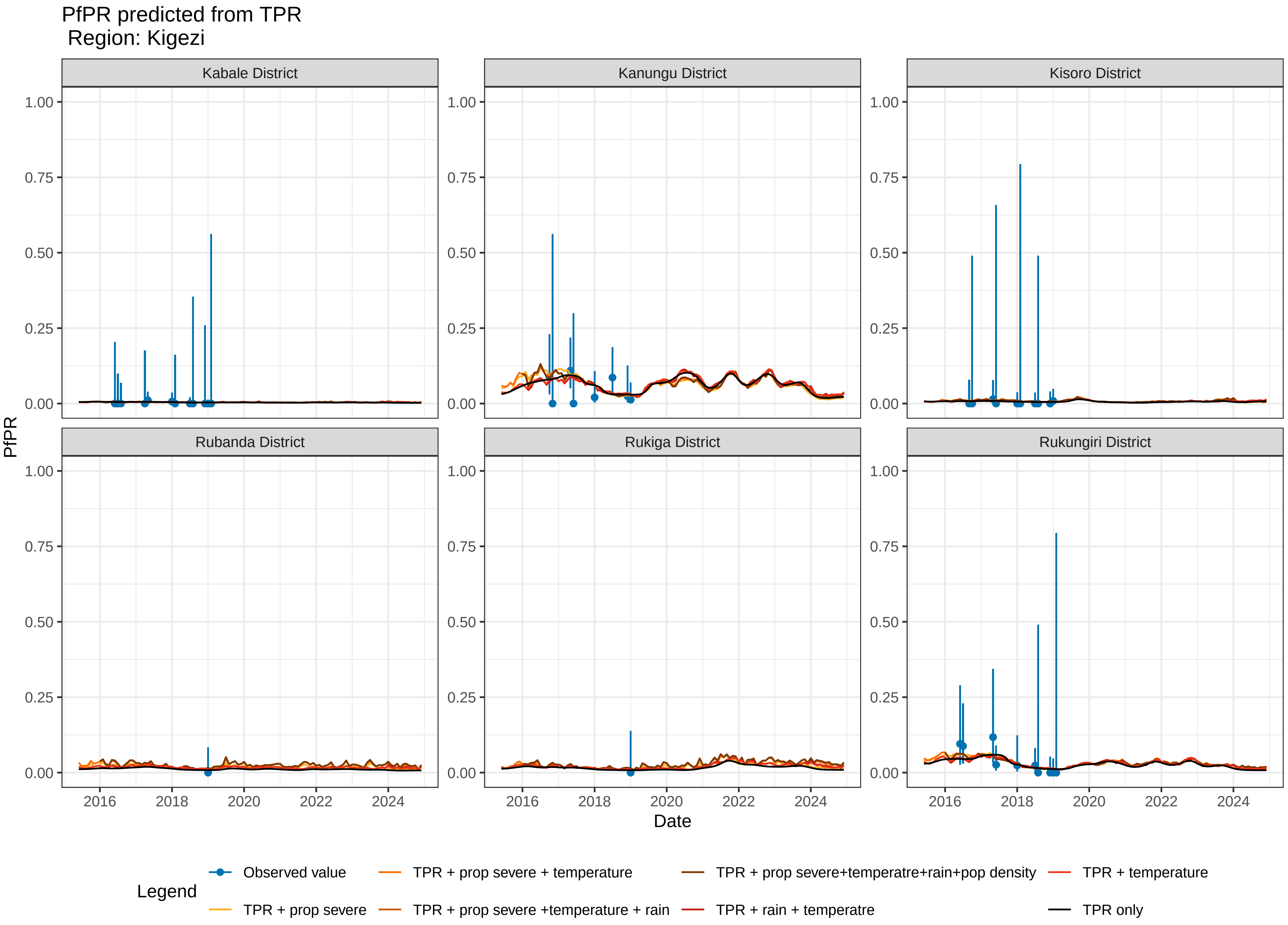
PfPR predicted from TPR
Region: Kigezi
Kabale District
Kanungu District
Kisoro District
Rubanda District
Rukiga District
Rukungiri District
PfPR
Date
1.00
0.75
0.50
0.25
0.00
2016
2018
2020
2022
2024
Legend
Observed value
TPR + prop severe
TPR + prop severe + temperature
TPR + prop severe +temperature + rain
TPR + prop severe+temperatre+rain+pop density
TPR + rain + temperatre
TPR + temperature
TPR only

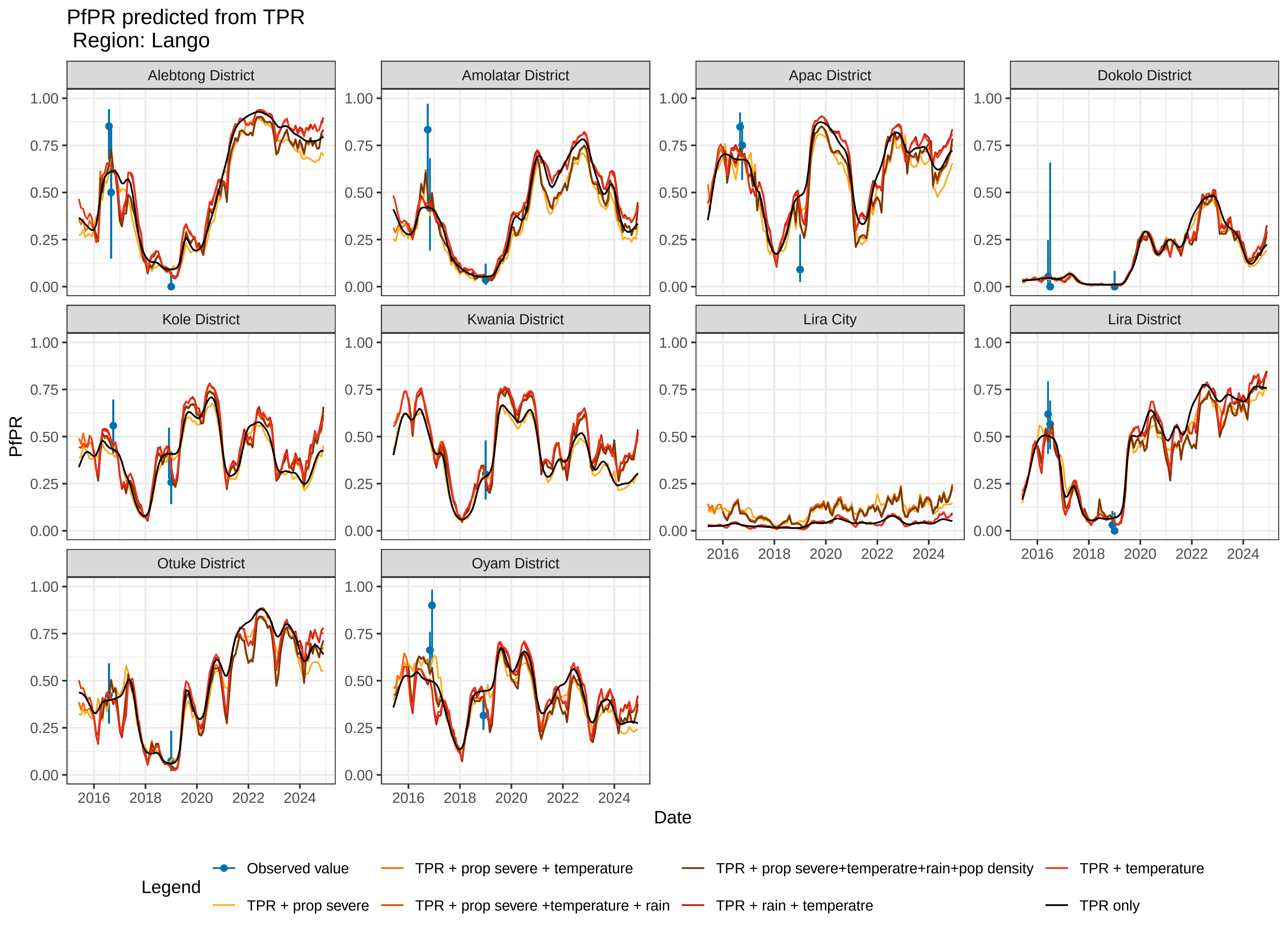

PfPR predicted from TPR
Region: Lango
Alebtong District
Amolatar District
Apac District
Dokolo District
Kole District
Kwania District
Lira City
Lira District
Otuke District
Oyam District
PfPR
Date
1.00
0.75
0.50
0.25
0.00
2016
2018
2020
2022
2024
Legend
Observed value
TPR + prop severe
TPR + prop severe + temperature
TPR + prop severe +temperature + rain
TPR + prop severe+temperatre+rain+pop density
TPR + rain + temperatre
TPR + temperature
TPR only

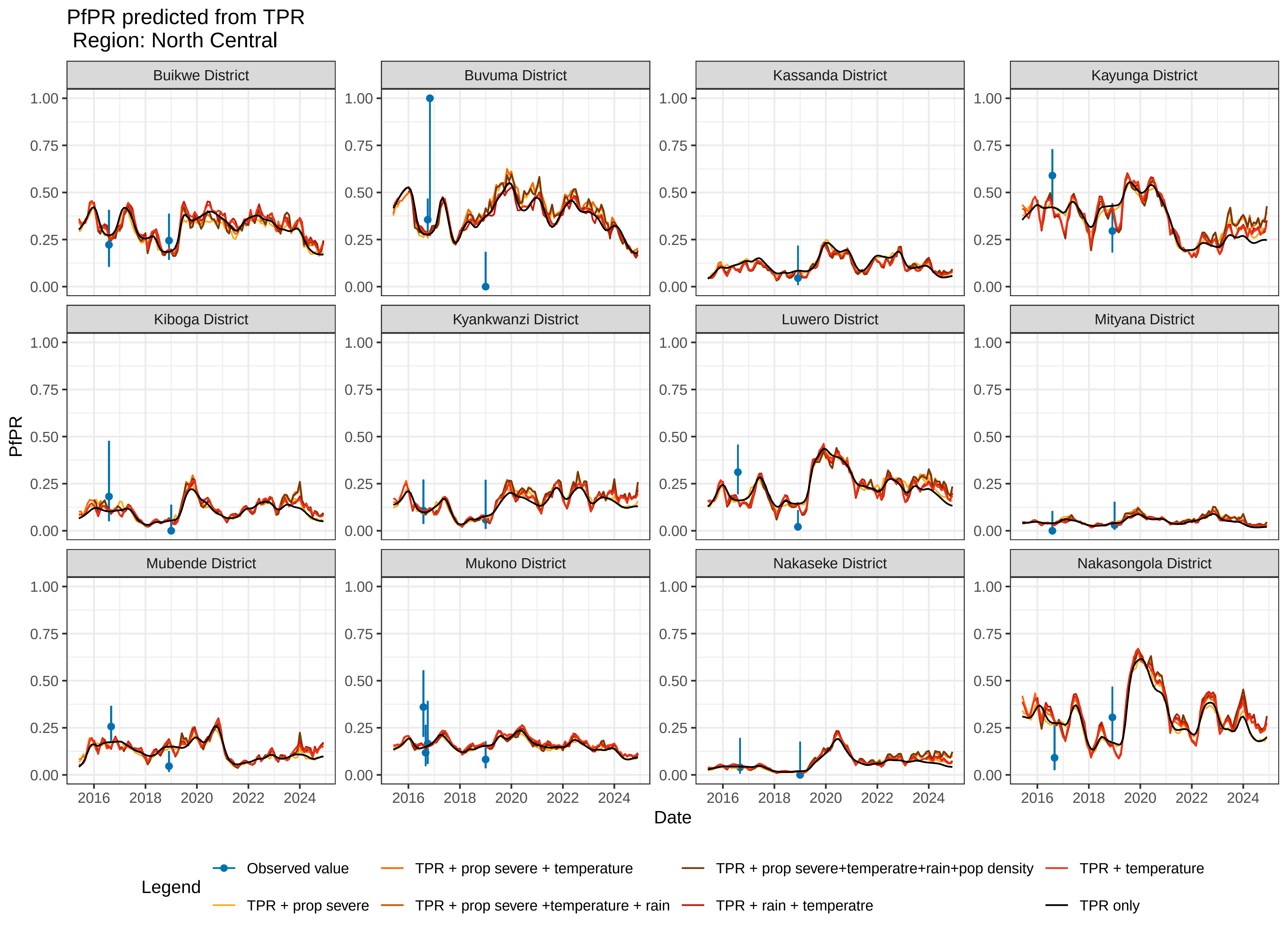

PfPR predicted from TPR
Region: North Central
Buikwe District
Buvuma District
Kassanda District
Kayunga District
Kiboga District
Kyankwanzi District
Luwero District
Mityana District
Mubende District
Mukono District
Nakaseke District
Nakasongola District
PfPR
0.00
0.25
0.50
0.75
1.00
2016
2018
2020
2022
2024
Date
Legend
Observed value
TPR + prop severe
TPR + prop severe + temperature
TPR + prop severe +temperature + rain
TPR + prop severe+temperatre+rain+pop density
TPR + rain + temperatre
TPR + temperature
TPR only

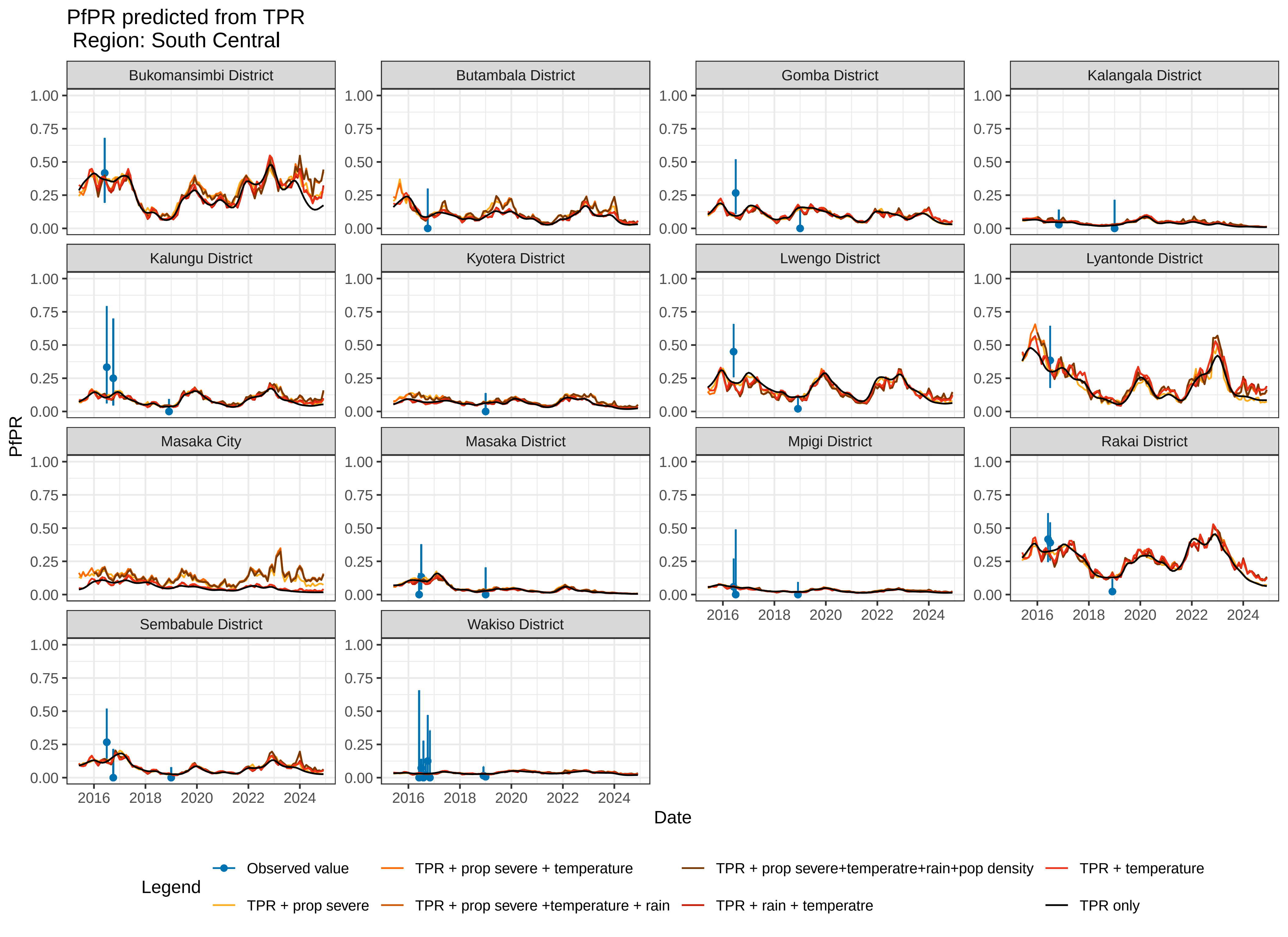
PfPR predicted from TPR
Region: South Central
Bukomansimbi District
Butambala District
Gomba District
Kalangala District
Kalungu District
Kyotera District
Lwengo District
Lyantonde District
Masaka City
Masaka District
Mpigi District
Rakai District
Sembabule District
Wakiso District
PfPR
0.00
0.25
0.50
0.75
1.00
Date
2016
2018
2020
2022
2024
Legend
Observed value
TPR + prop severe
TPR + prop severe + temperature
TPR + prop severe +temperature + rain
TPR + prop severe+temperatre+rain+pop density
TPR + rain + temperatre
TPR + temperature
TPR only

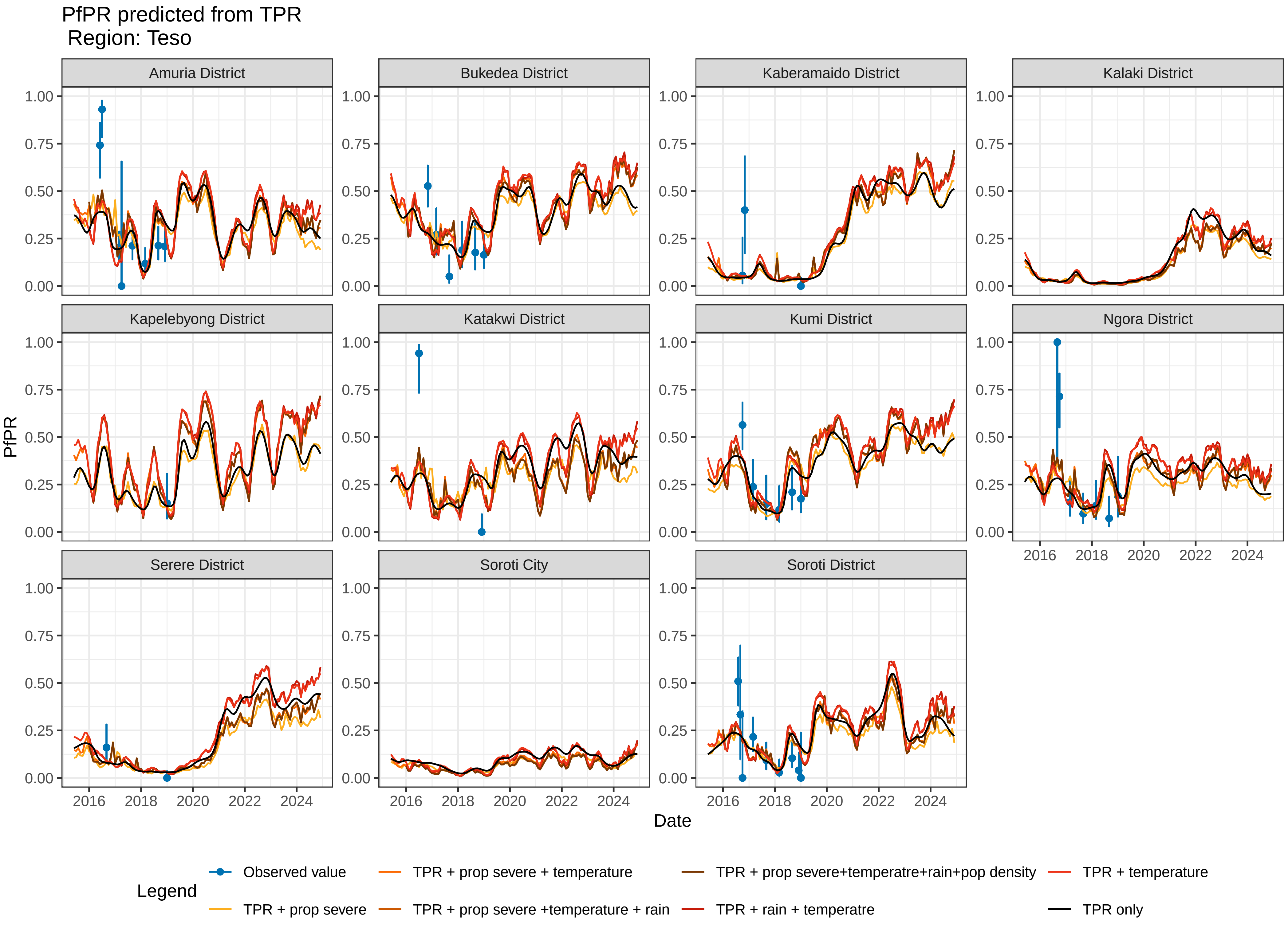
PfPR predicted from TPR
Region: Teso
Amuria District
Bukedea District
Kaberamaido District
Kalaki District
Kapelebyong District
Katakwi District
Kumi District
Ngora District
Serere District
Soroti City
Soroti District
PfPR
Date
Legend
Observed value
TPR + prop severe
TPR + prop severe + temperature
TPR + prop severe +temperature + rain
TPR + prop severe+temperatre+rain+pop density
TPR + rain + temperatre
TPR + temperature
TPR only

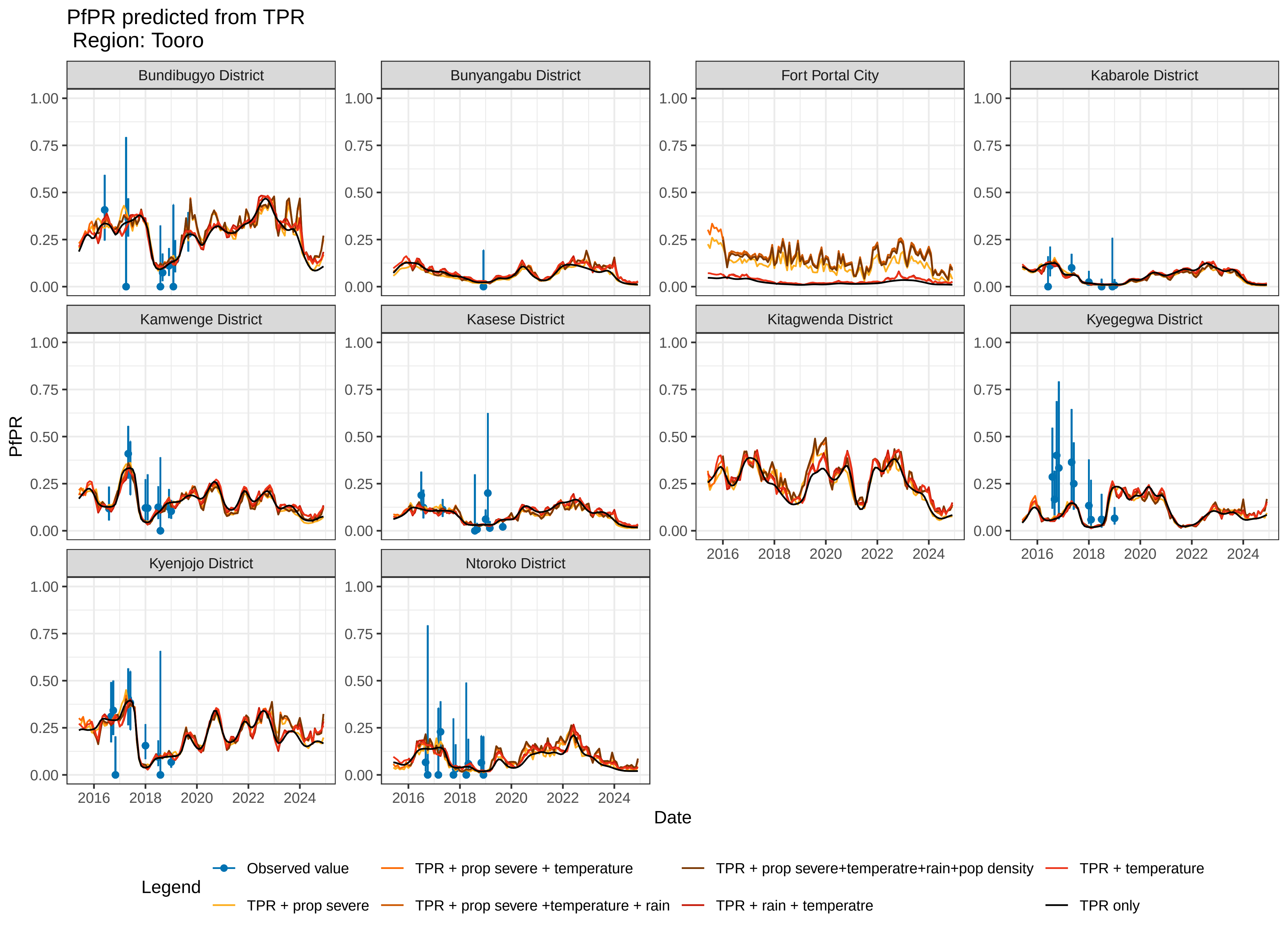
PfPR predicted from TPR
Region: Tooro
Bundibugyo District
Bunyangabu District
Fort Portal City
Kabarole District
Kamwenge District
Kasese District
Kitagwenda District
Kyegegwa District
Kyenjojo District
Ntoroko District
PfPR
0.00
0.25
0.50
0.75
1.00
2016
2018
2020
2022
2024
Date
Legend
Observed value
TPR + prop severe
TPR + prop severe + temperature
TPR + prop severe +temperature + rain
TPR + prop severe+temperatre+rain+pop density
TPR + rain + temperatre
TPR + temperature
TPR only

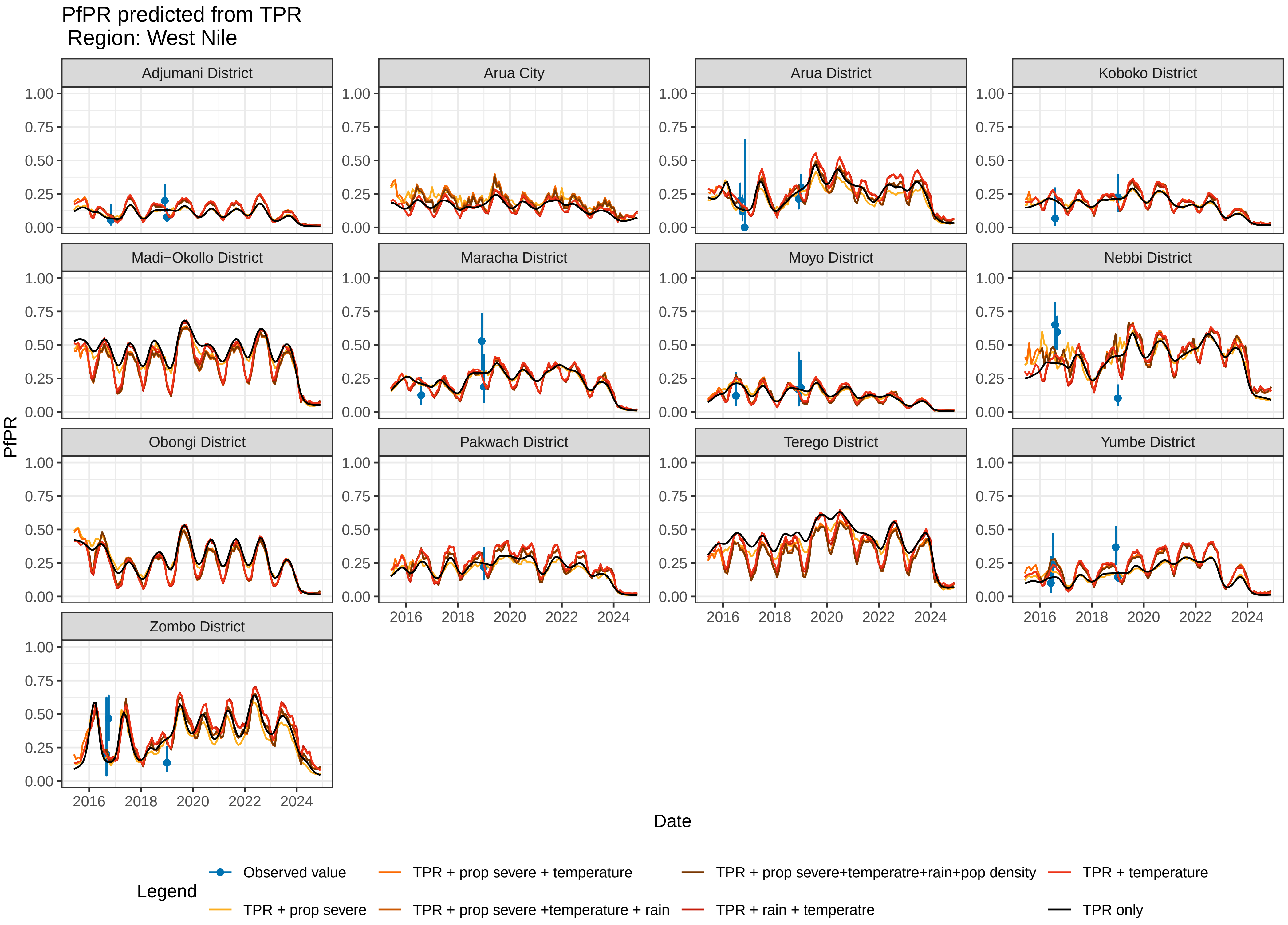

PfPR predicted from TPR
Region: West Nile
Adjumani District
Arua City
Arua District
Koboko District
Madi−Okollo District
Maracha District
Moyo District
Nebbi District
Obongi District
Pakwach District
Terego District
Yumbe District
Zombo District
PfPR
0.00
0.25
0.50
0.75
1.00
Date
2016
2018
2020
2022
2024
Legend
Observed value
TPR + prop severe
TPR + prop severe + temperature
TPR + prop severe +temperature + rain
TPR + prop severe+temperatre+rain+pop density
TPR + rain + temperatre
TPR + temperature
TPR only
