## Supplemental Figure 2 for "Estimating *Plasmodium falciparum* Parasite Rate using Test Positivity Rate from 2016-2024: Health Management Information Systems in Uganda"

West Nile − Page 1

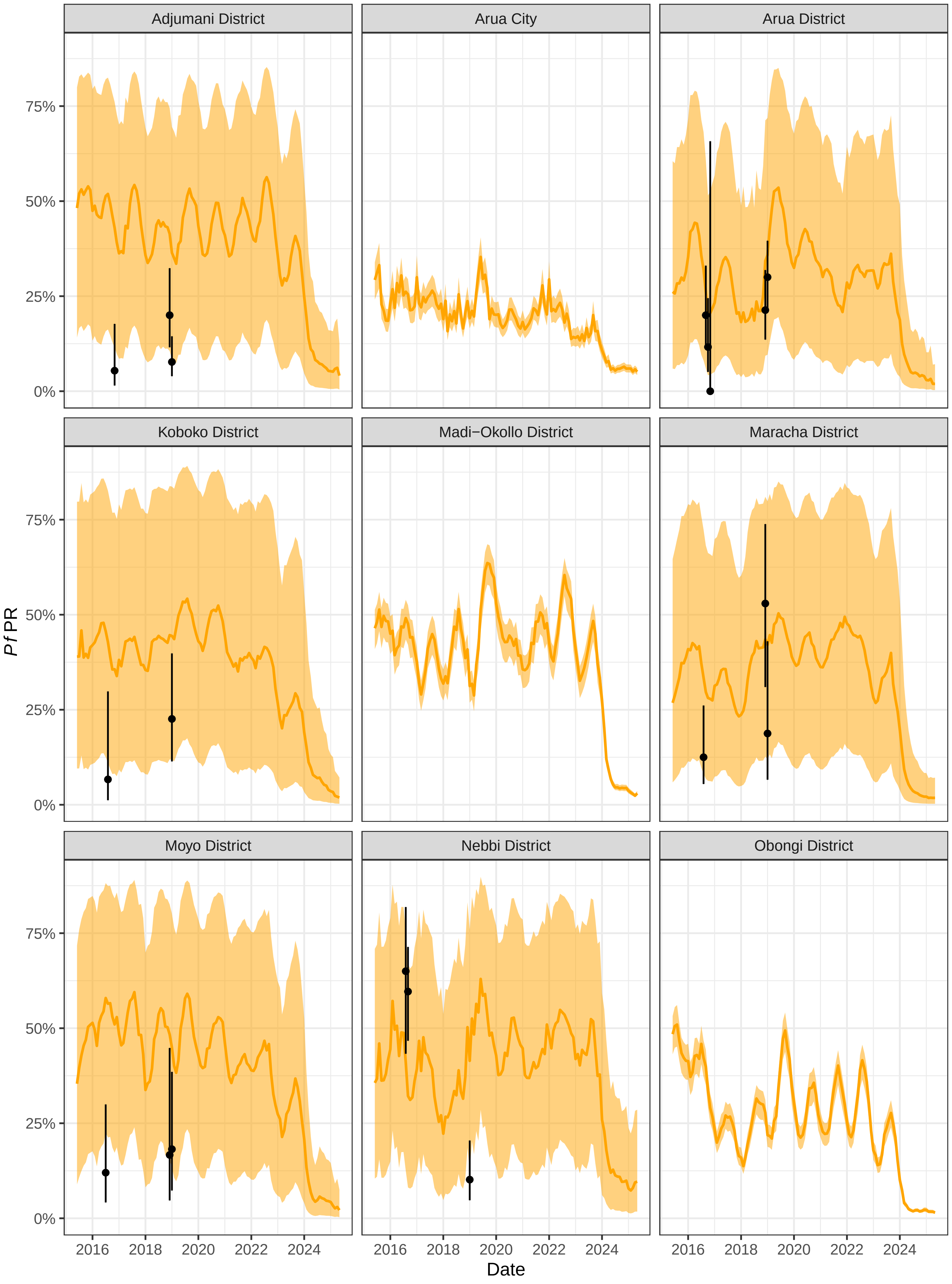

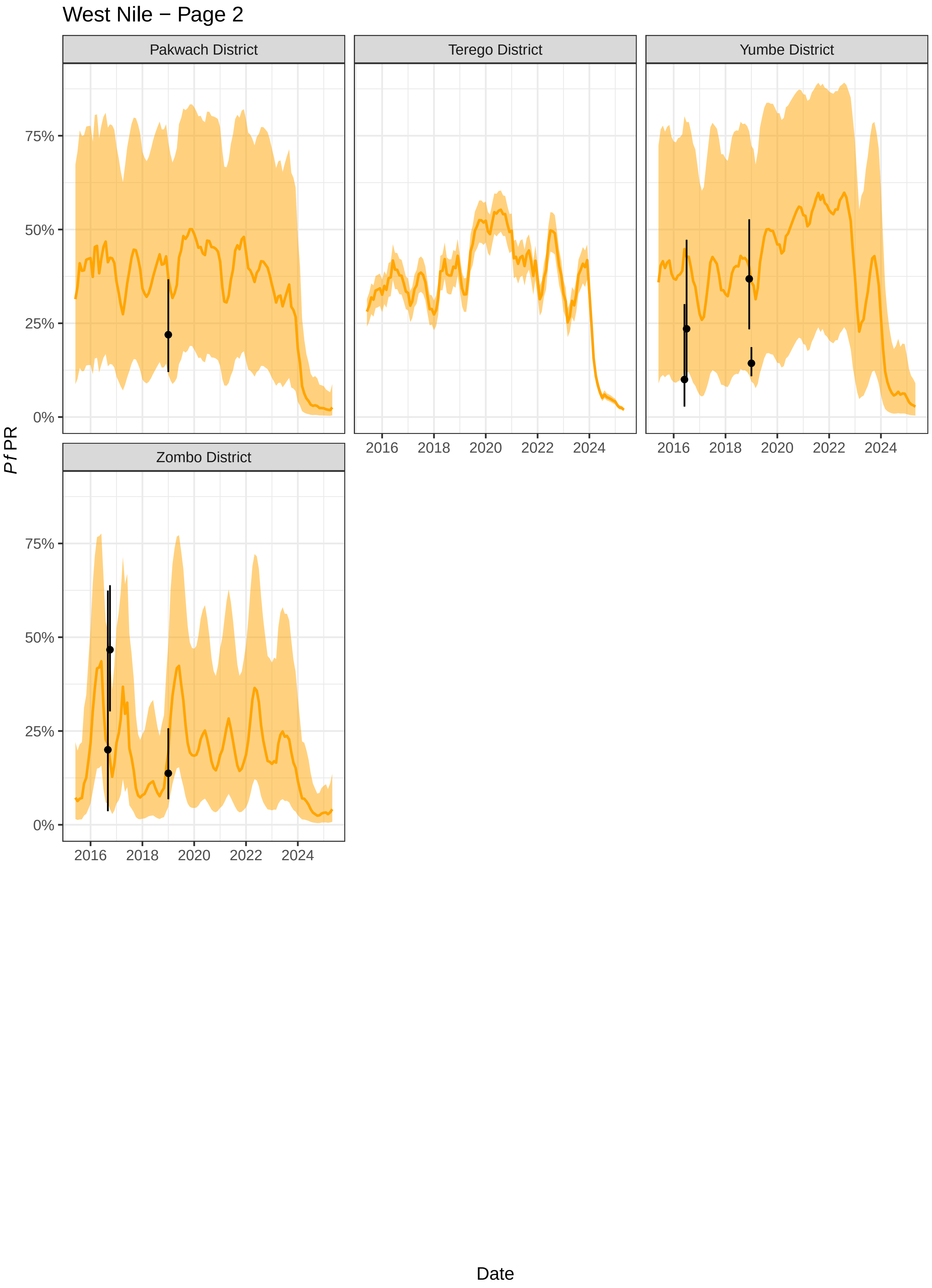
West Nile − Page 2
Pakwach District
Terego District
Yumbe District
Zombo District
75%
50%
25%
0%
2016
2018
2020
2022
2024
Pf PR
Date

### Karamoja − Page 1

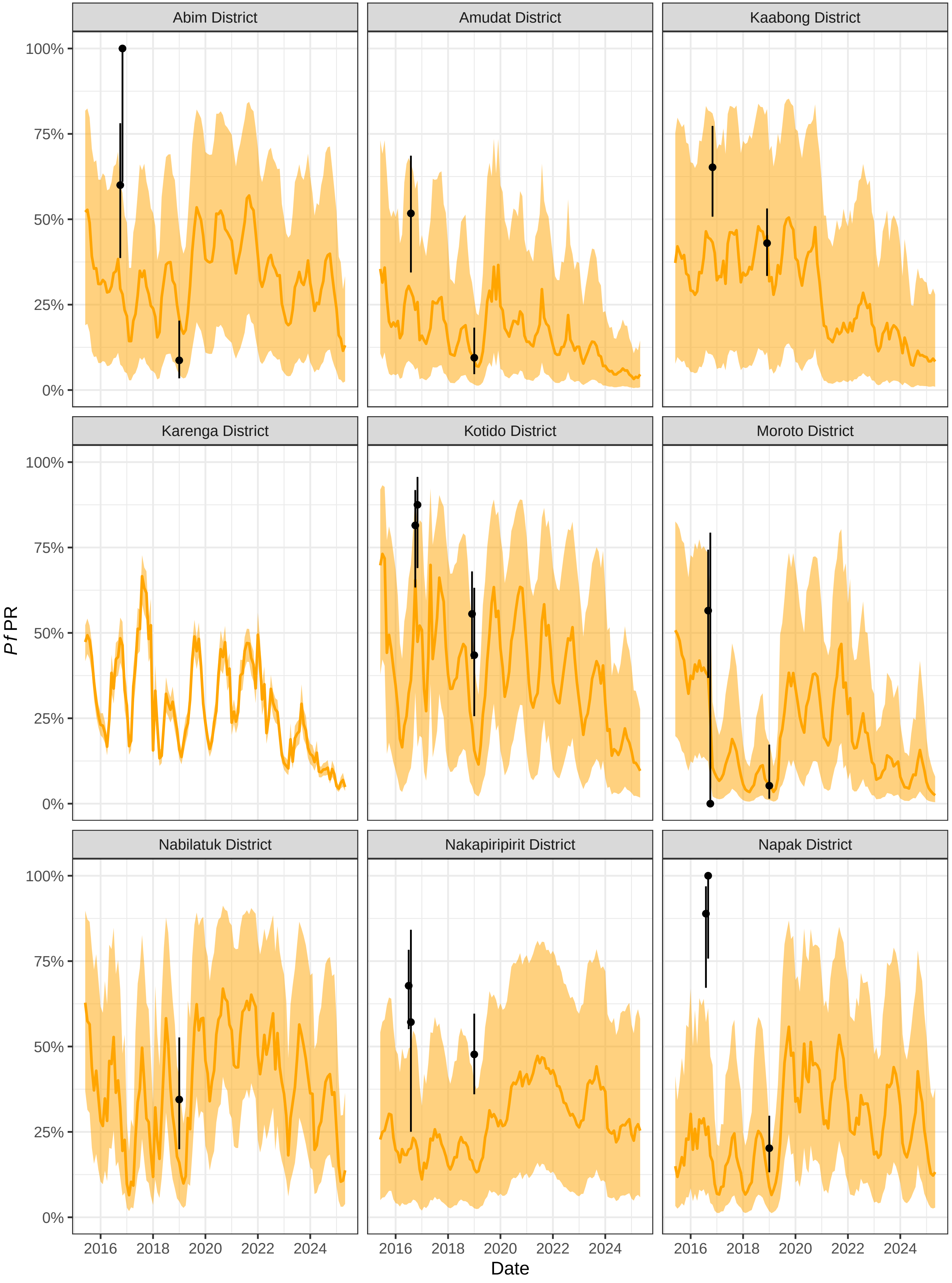

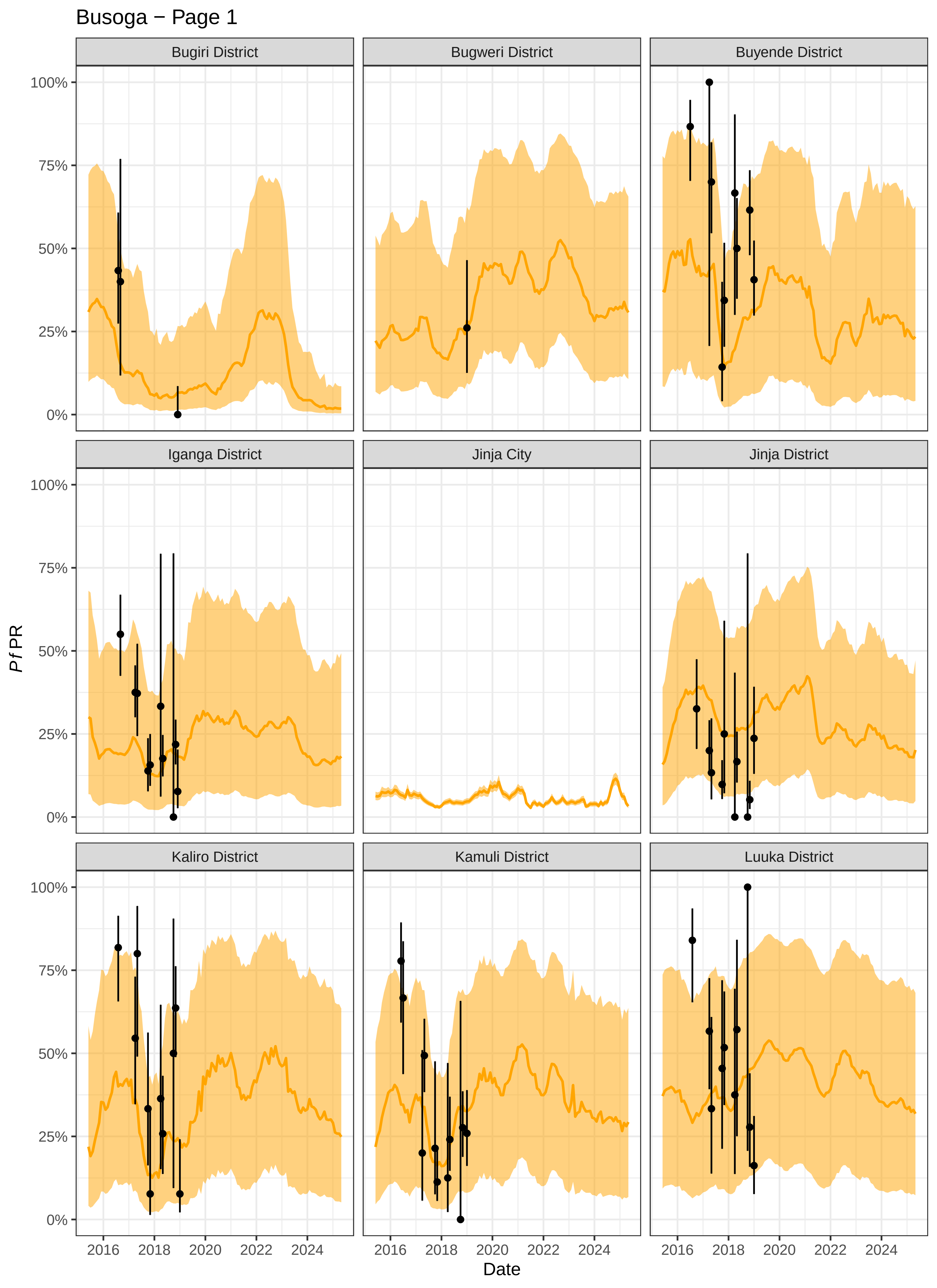

Busoga − Page 1
Bugiri District
Bugweri District
Buyende District
Iganga District
Jinja City
Jinja District
Kaliro District
Kamuli District
Luuka District
100%
75%
50%
25%
0%
*Pf*PR
2016
2018
2020
2022
2024
Date

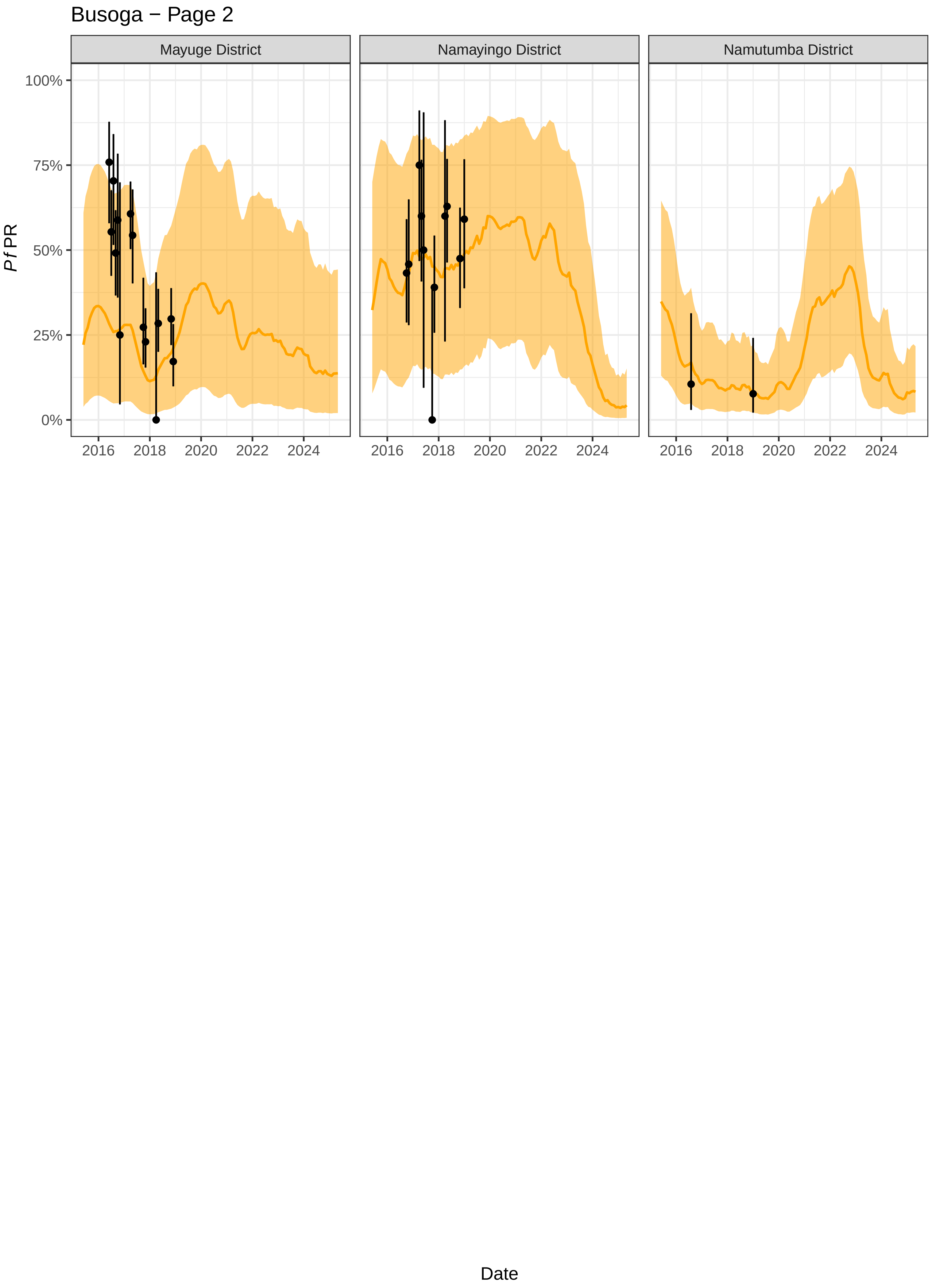

Busoga − Page 2
Mayuge District
Namayingo District
Namutumba District
100%
75%
50%
25%
0%
Pf PR
2016
2018
2020
2022
2024
Date

### Bugisu − Page 1

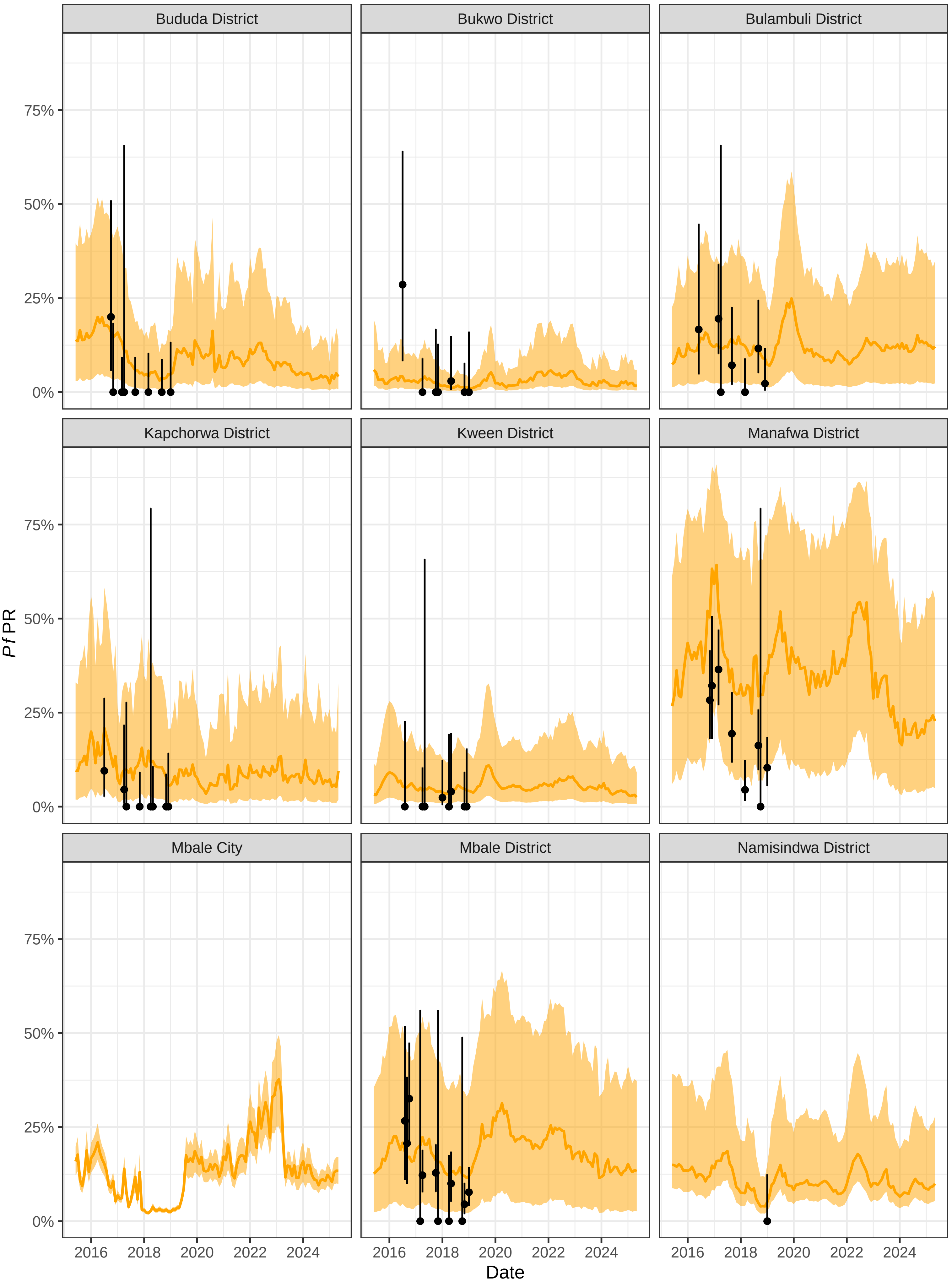

Bugisu − Page 2

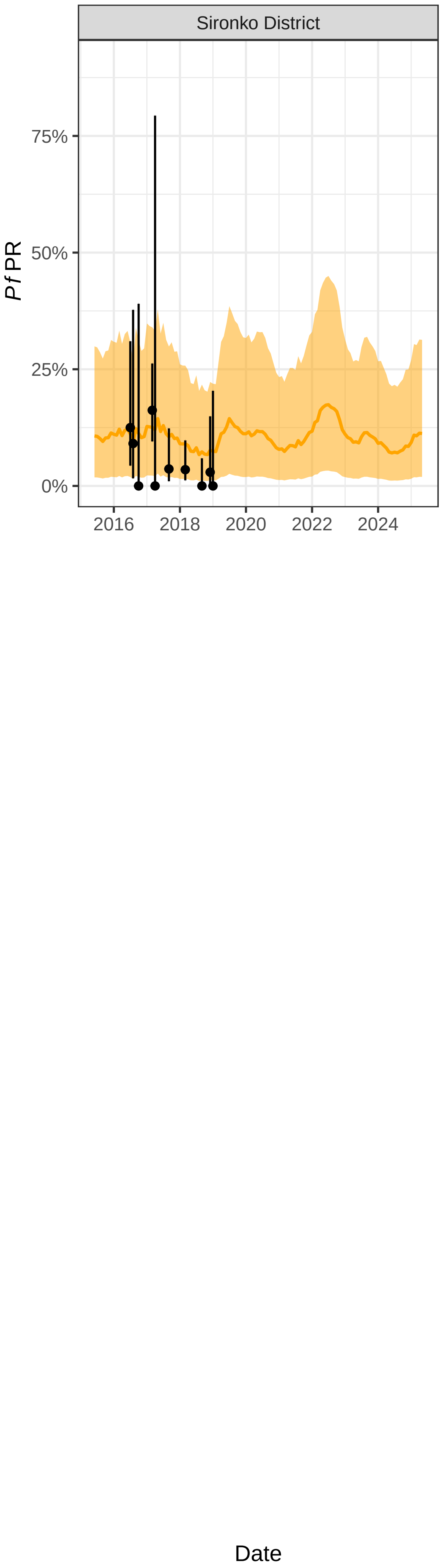

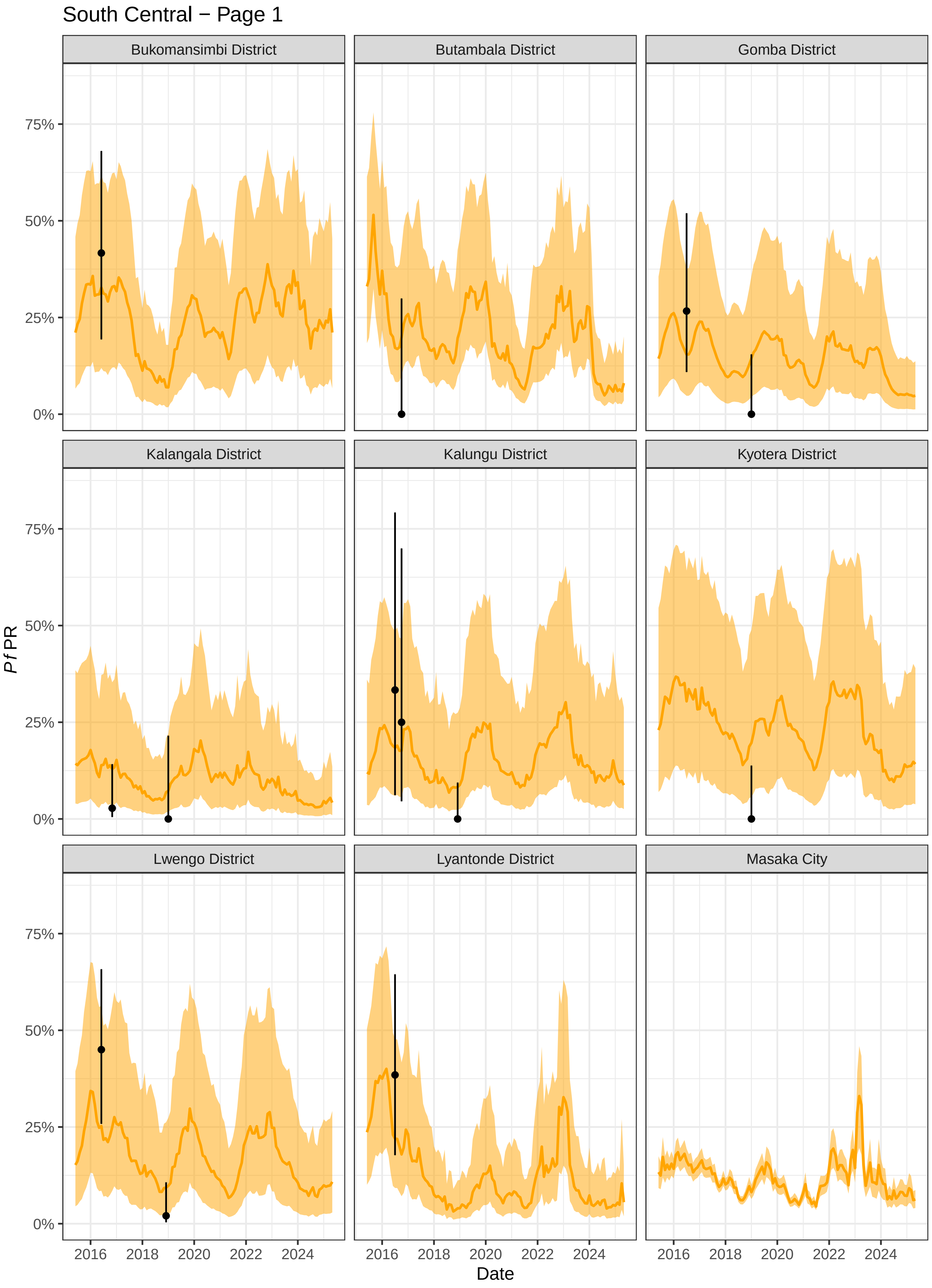
South Central − Page 1
Bukomansimbi District
Butambala District
Gomba District
Kalangala District
Kalungu District
Kyotera District
Lwengo District
Lyantonde District
Masaka City
Pf PR
75%
50%
25%
0%
2016
2018
2020
2022
2024
Date

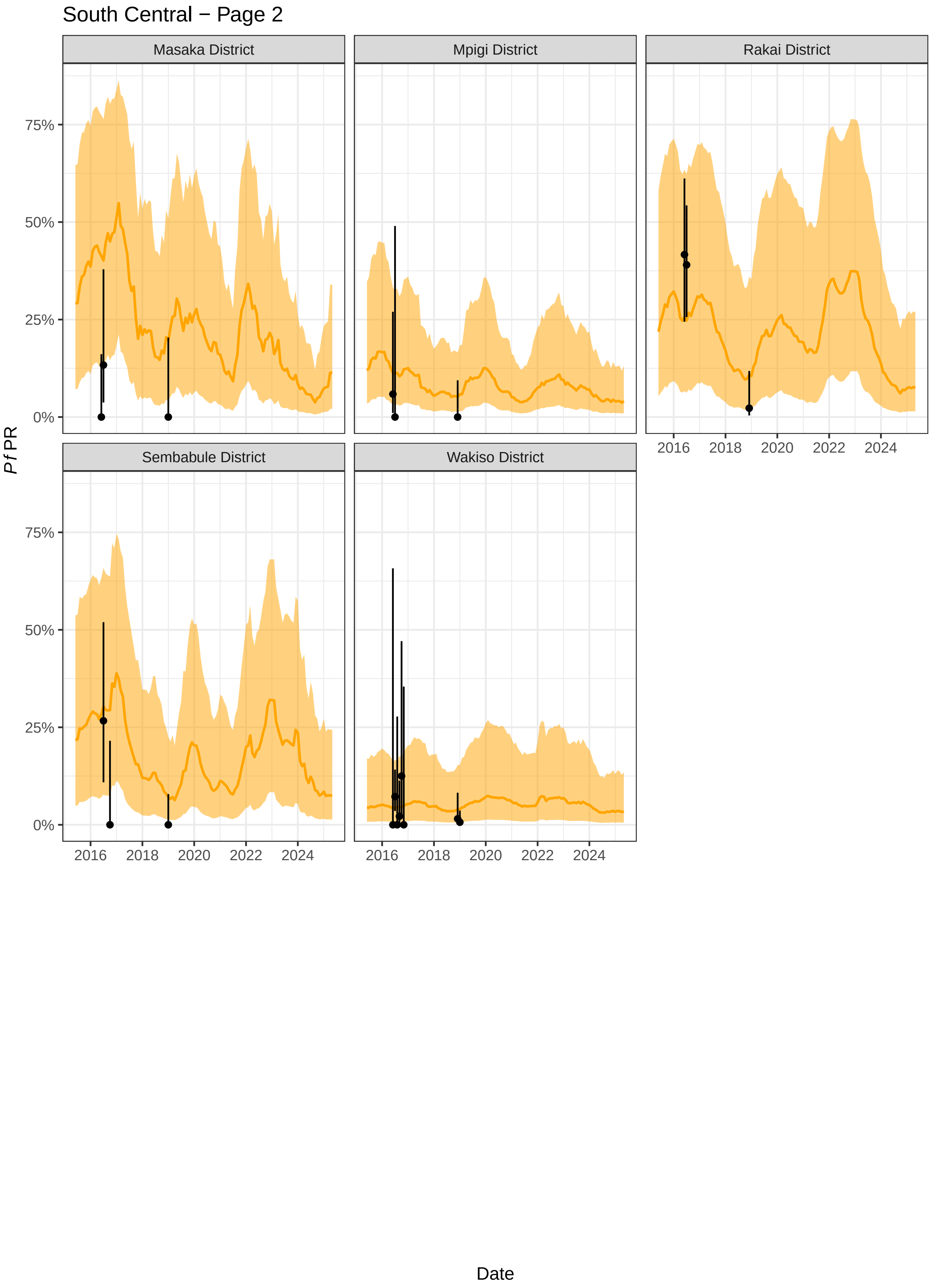
South Central − Page 2
Masaka District
Mpigi District
Rakai District
Sembabule District
Wakiso District
0%
25%
50%
75%
2016
2018
2020
2022
2024
Pf PR
Date

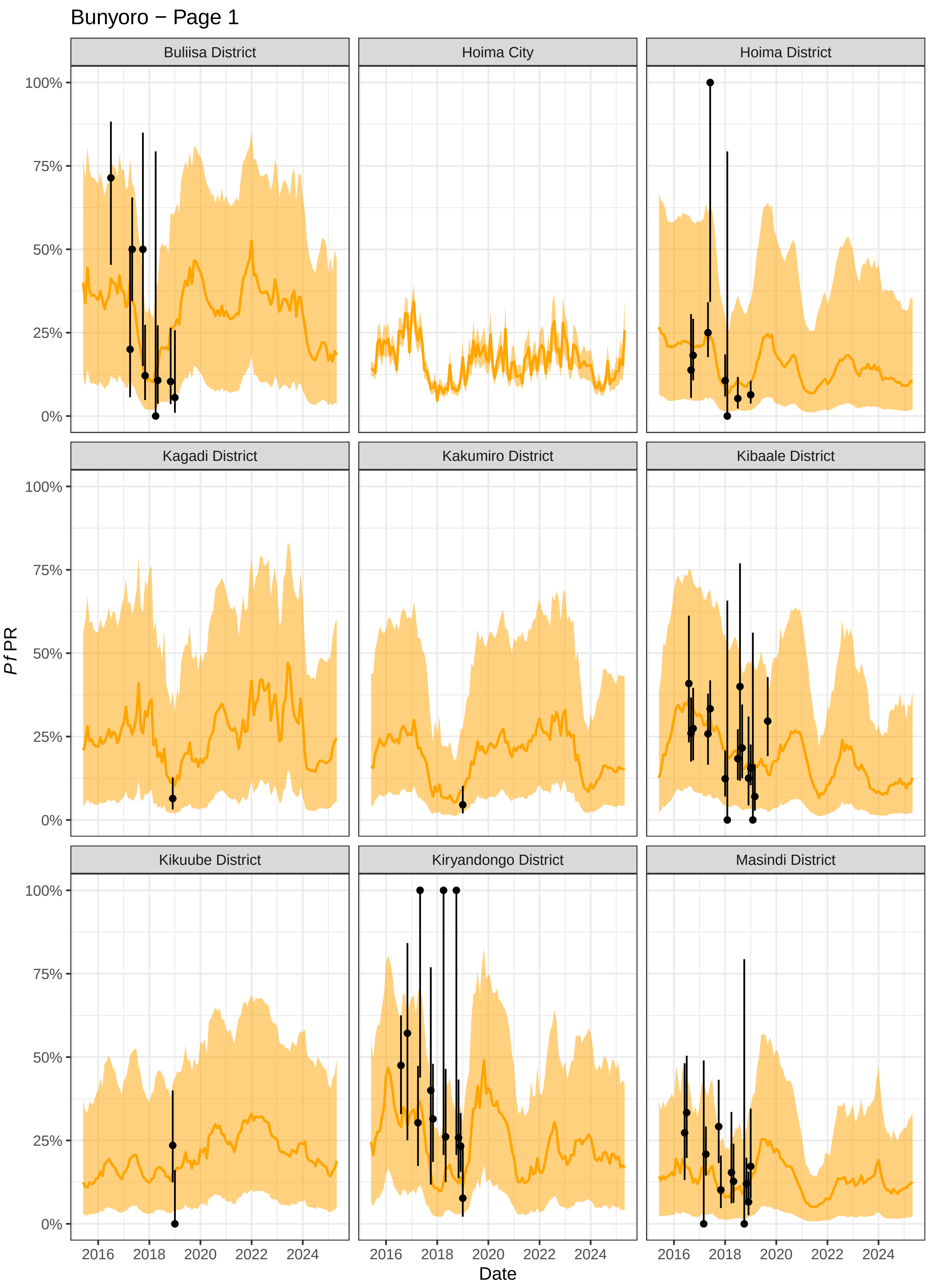
Bunyoro − Page 1
Buliisa District
Hoima City
Hoima District
Kagadi District
Kakumiro District
Kibaale District
Kikuube District
Kiryandongo District
Masindi District
100%
75%
50%
25%
0%
2016
2018
2020
2022
2024
Pf PR
Date

Acholi − Page 1

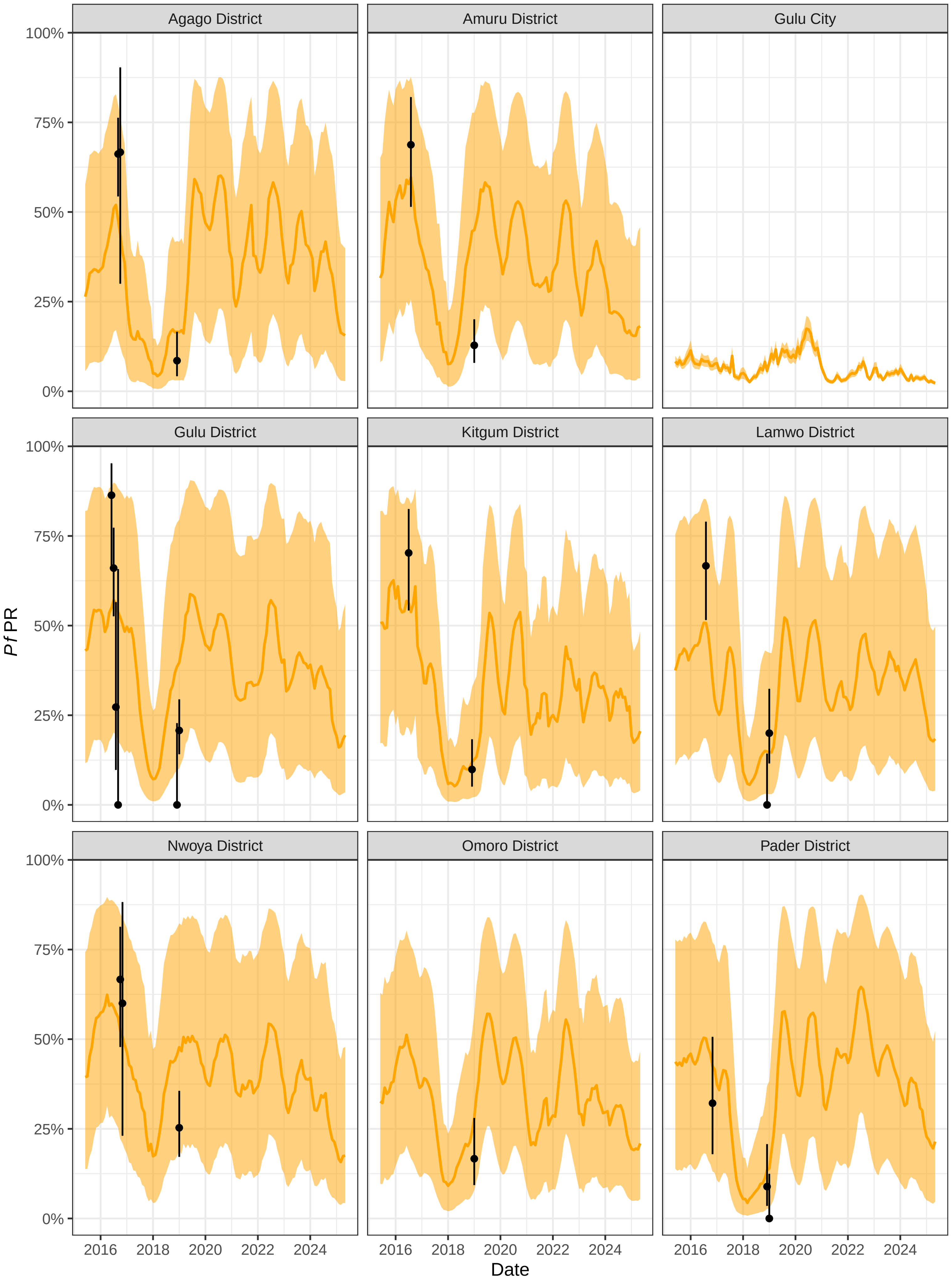

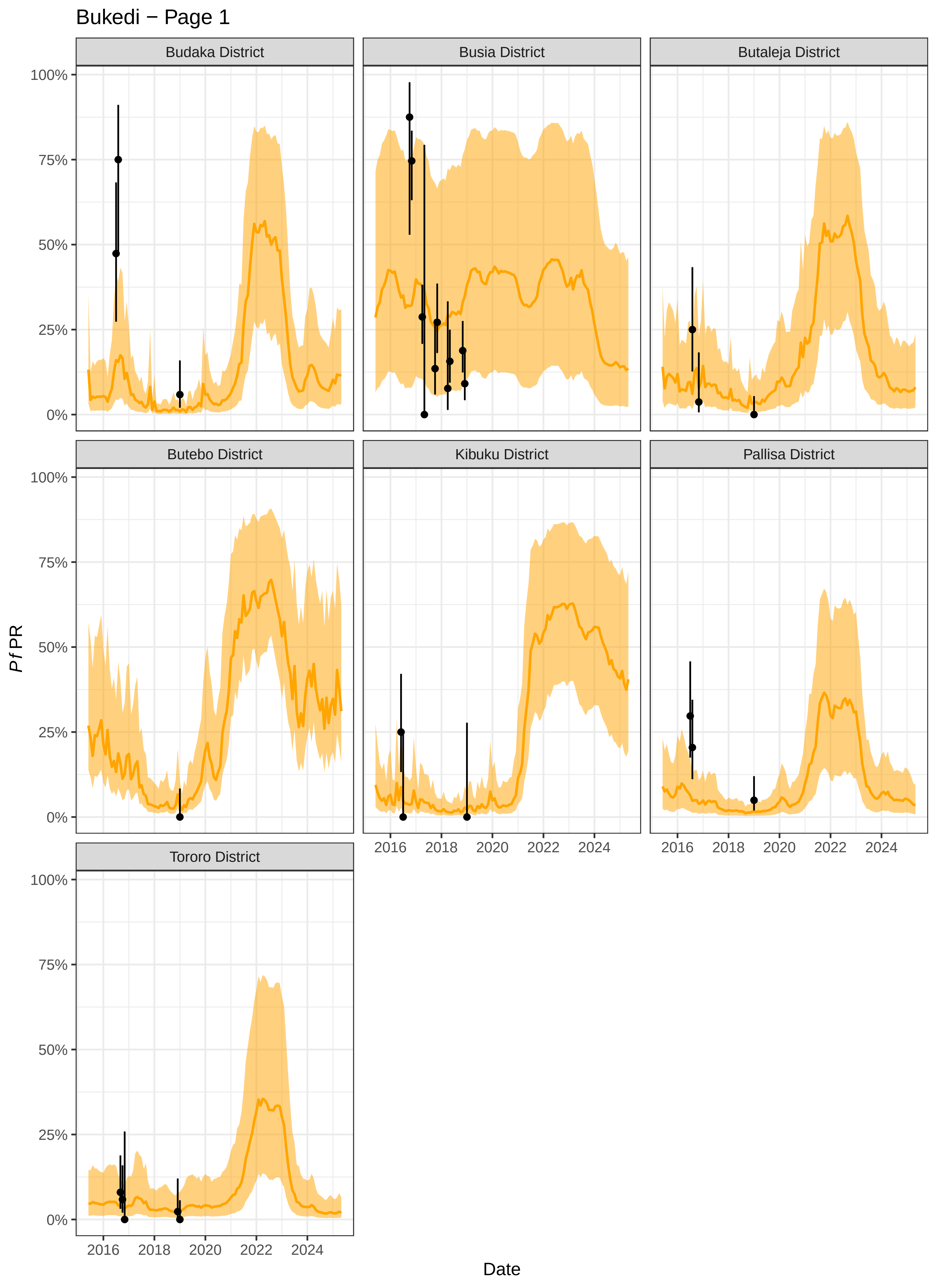
Bukedi − Page 1
Budaka District
Busia District
Butaleja District
Butebo District
Kibuku District
Pallisa District
Tororo District
100%
75%
50%
25%
0%
2016
2018
2020
2022
2024
*Pf*PR
Date

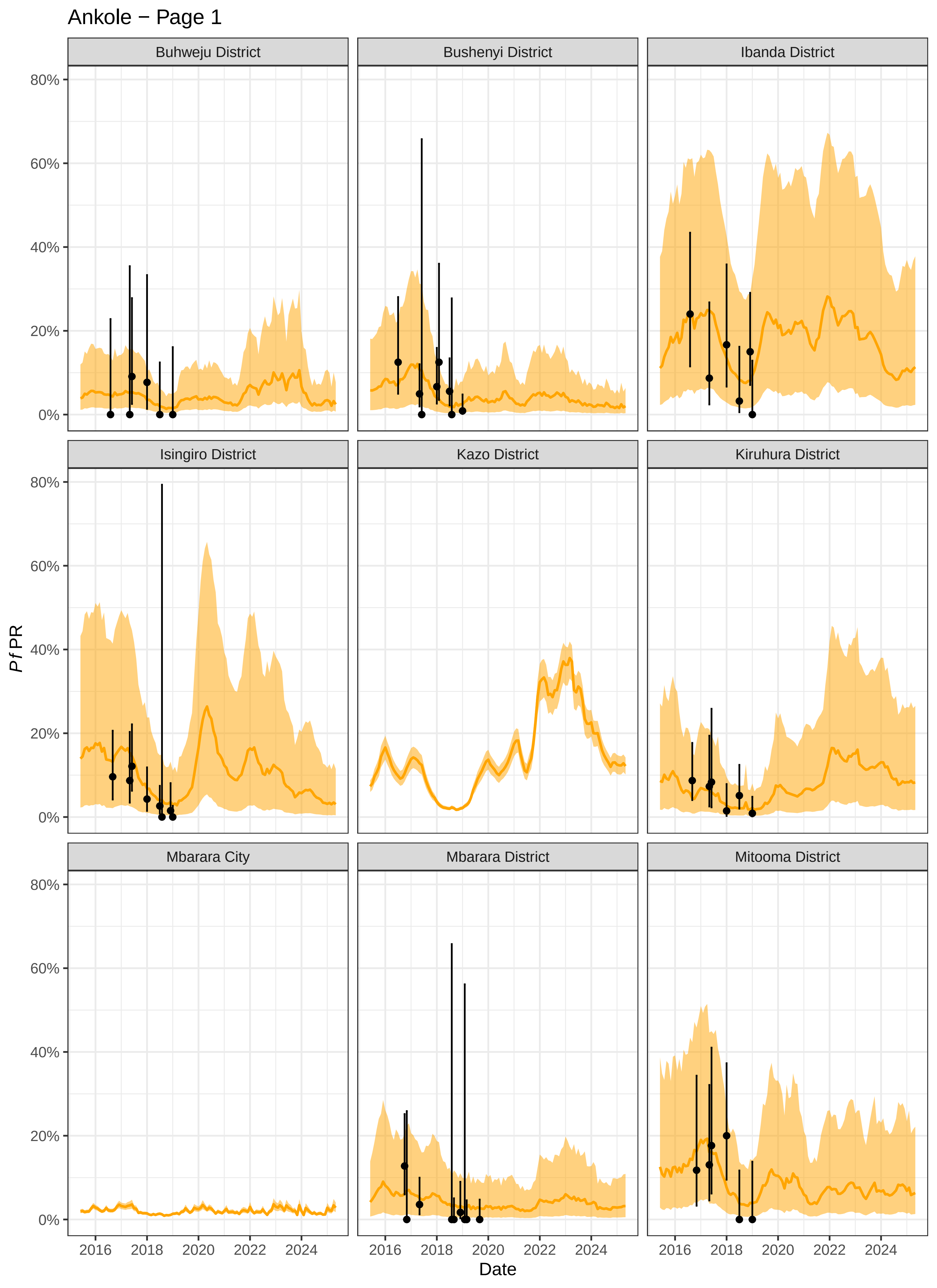
Ankole − Page 1
Buhweju District
Bushenyi District
Ibanda District
Isingiro District
Kazo District
Kiruhura District
Mbarara City
Mbarara District
Mitooma District
0%
20%
40%
60%
80%
2016
2018
2020
2022
2024
Pf PR
Date

#### Ankole − Page 2

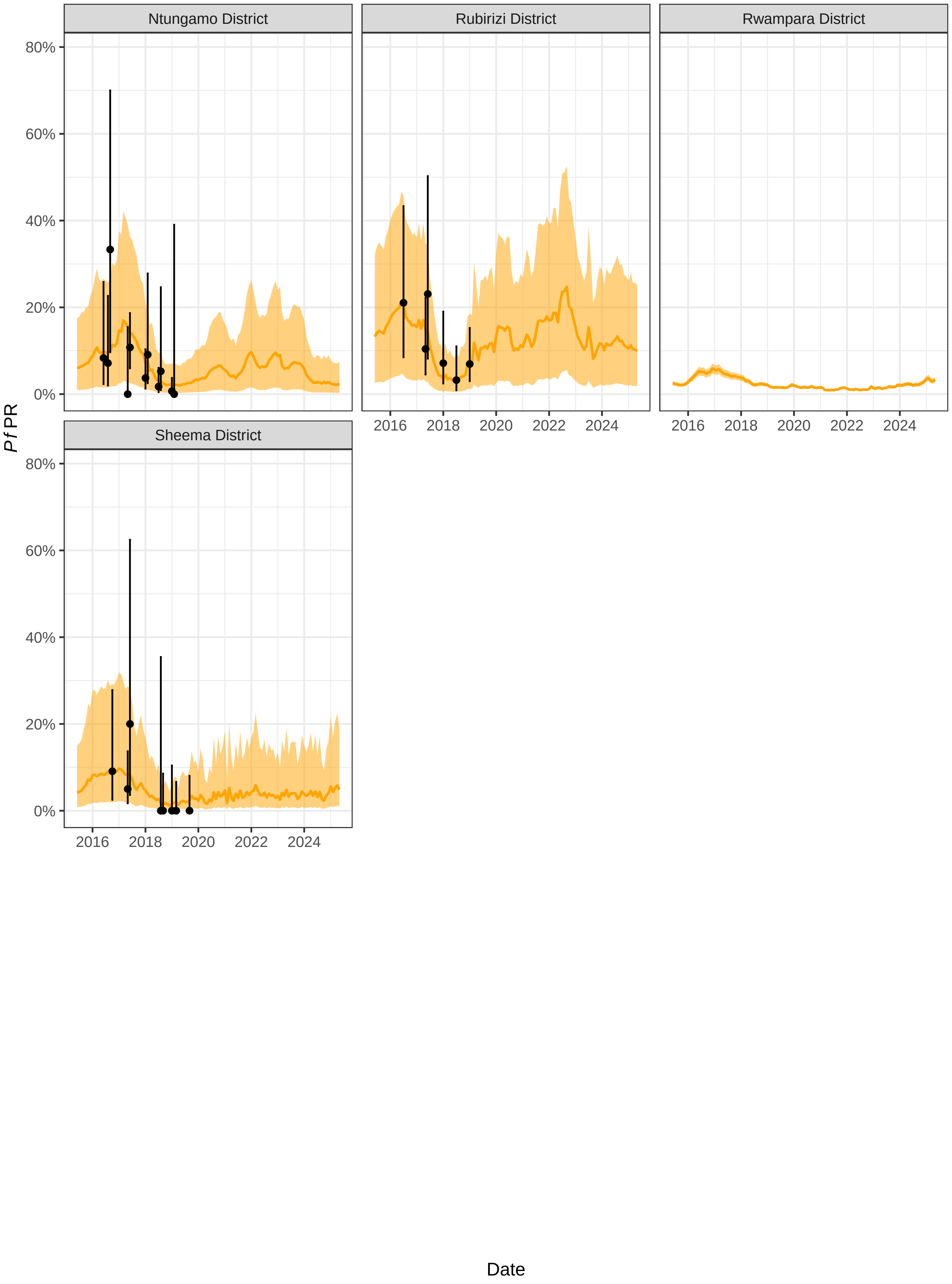

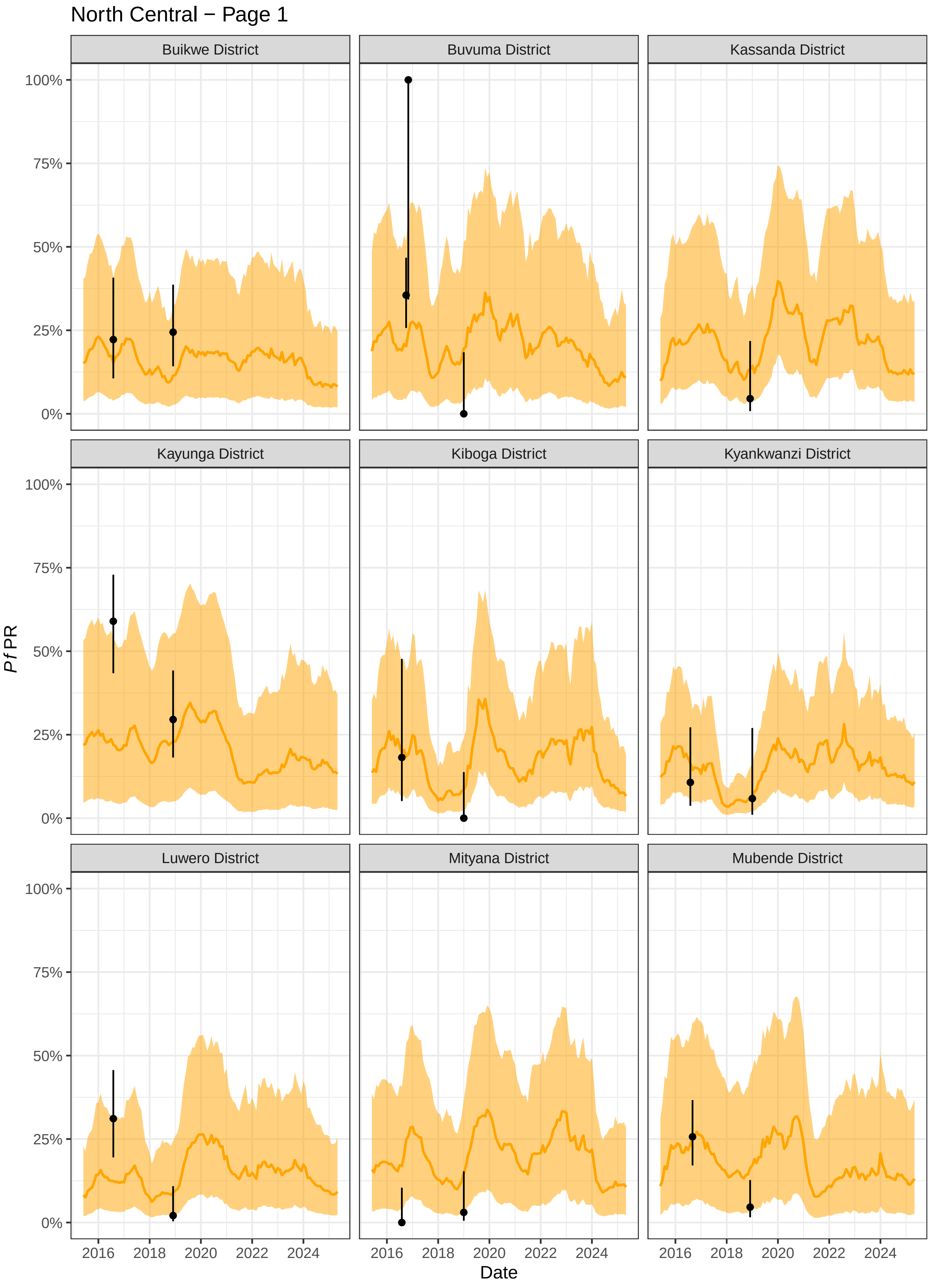

North Central − Page 1
Buikwe District
Buvuma District
Kassanda District
Kayunga District
Kiboga District
Kyankwanzi District
Luwero District
Mityana District
Mubende District
100%
75%
50%
25%
0%
Pf PR
2016
2018
2020
2022
2024
Date

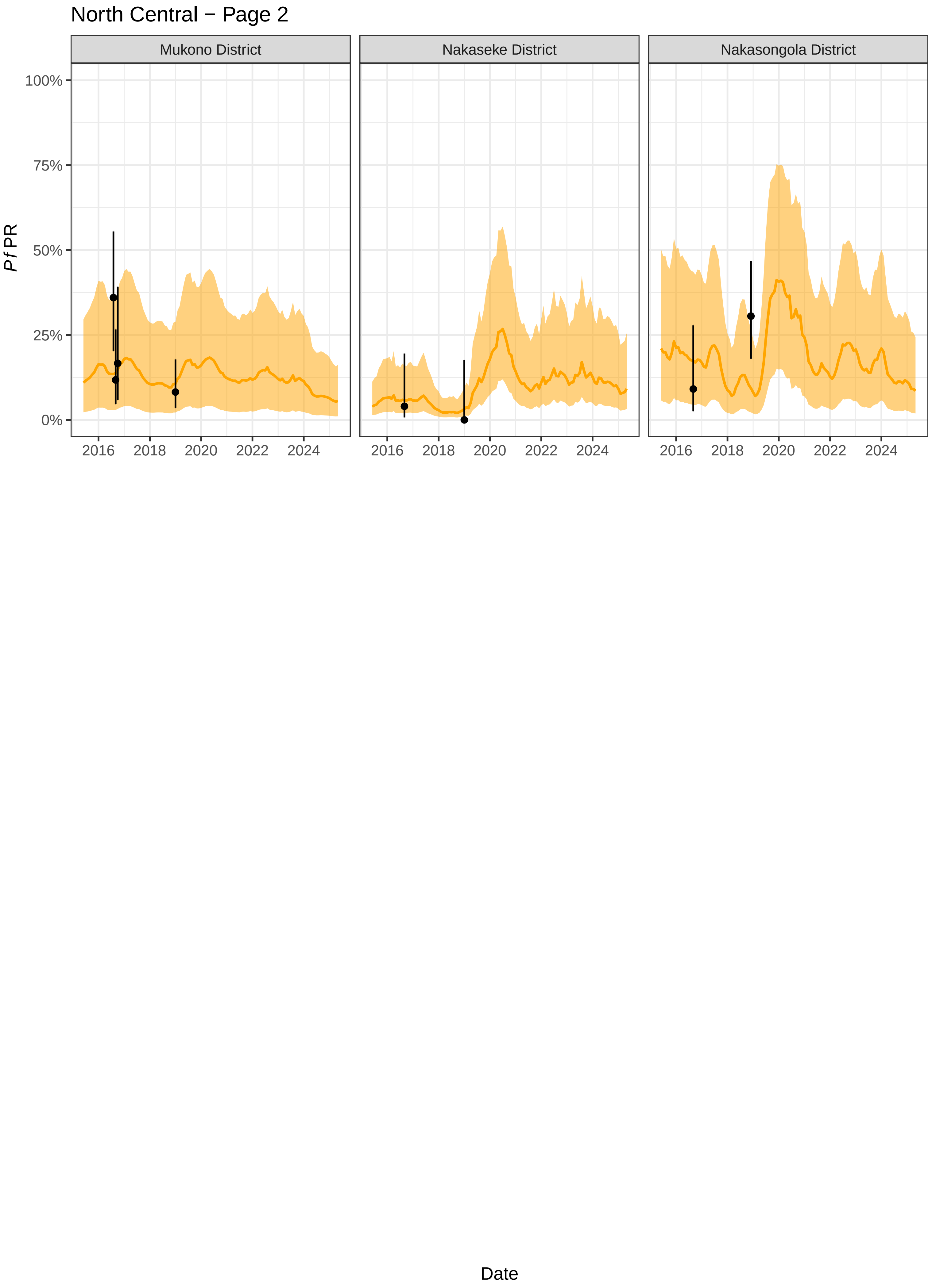
North Central − Page 2
Mukono District
Nakaseke District
Nakasongola District
100%
75%
50%
25%
0%
*Pf*PR
2016
2018
2020
2022
2024
Date

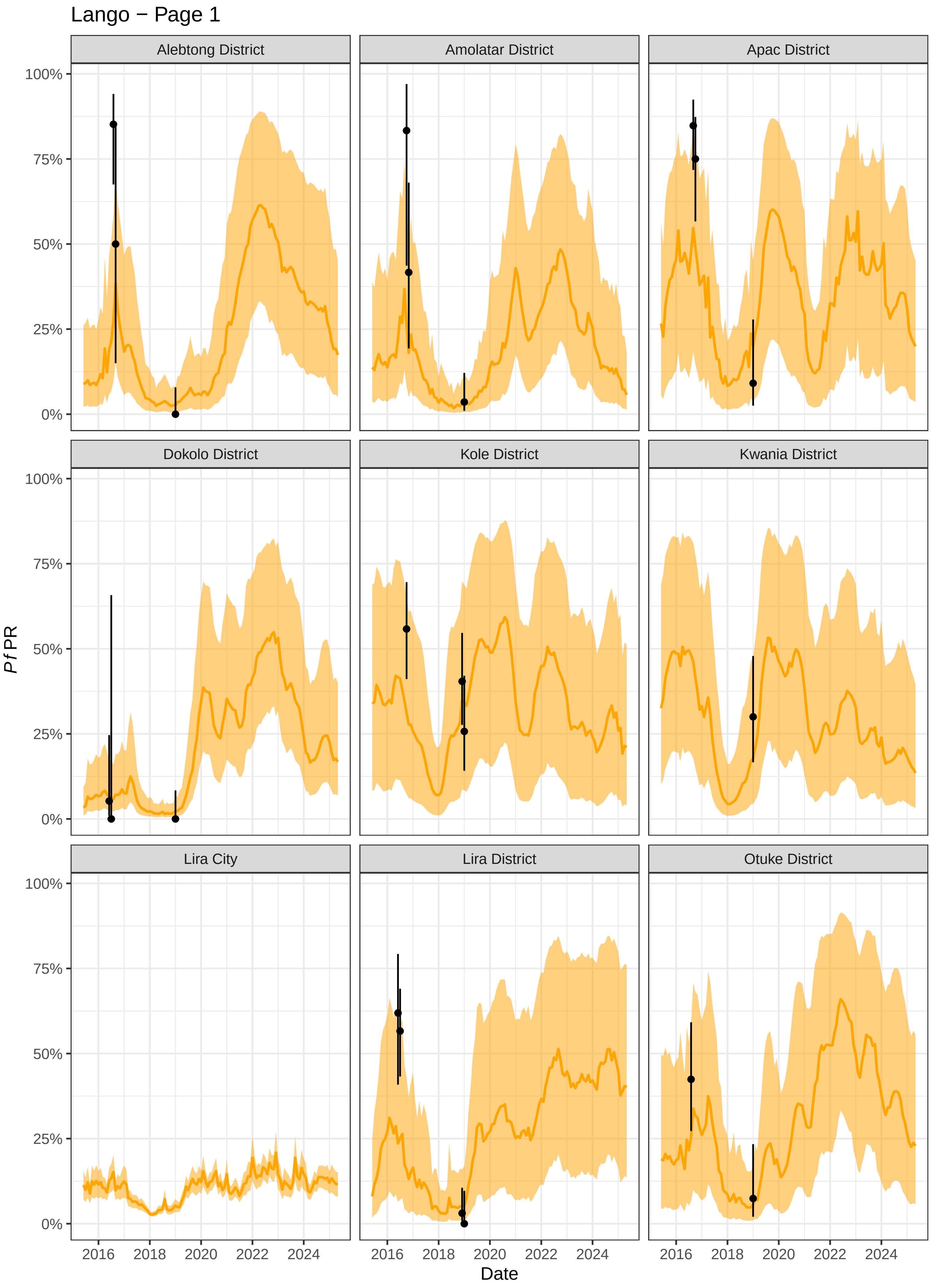
Lango − Page 1
Alebtong District
Amolatar District
Apac District
Dokolo District
Kole District
Kwania District
Lira City
Lira District
Otuke District
100%
75%
50%
25%
0%
Pf PR
2016
2018
2020
2022
2024
Date

#### Lango − Page 2

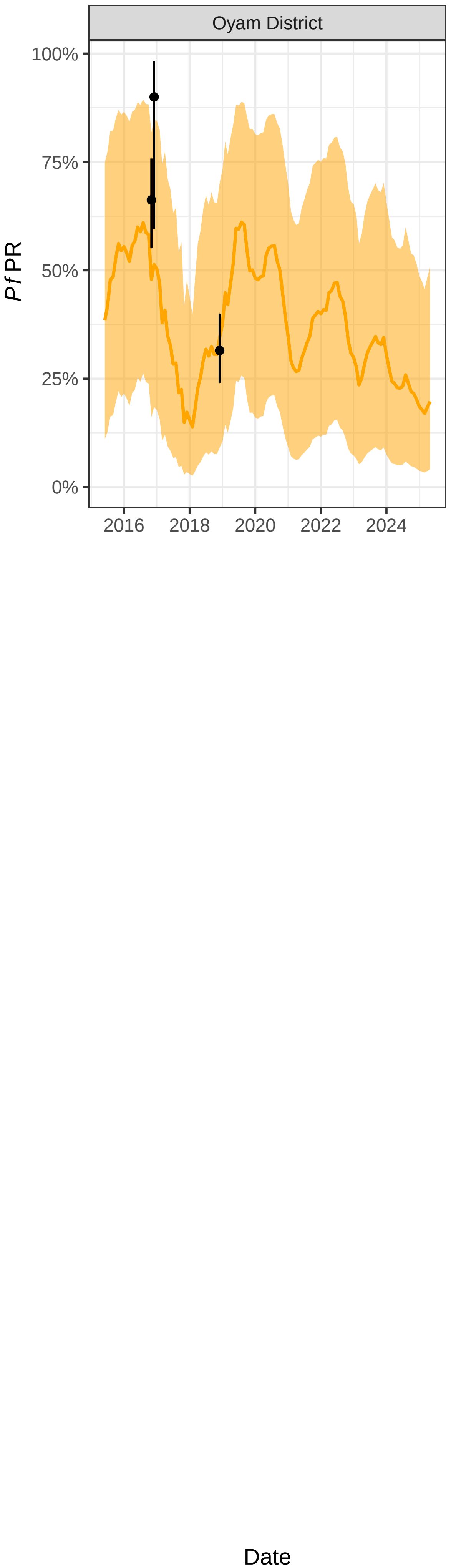

Oyam District
100%
75%
50%
25%
0%
PfPR
2016
2018
2020
2022
2024
Date

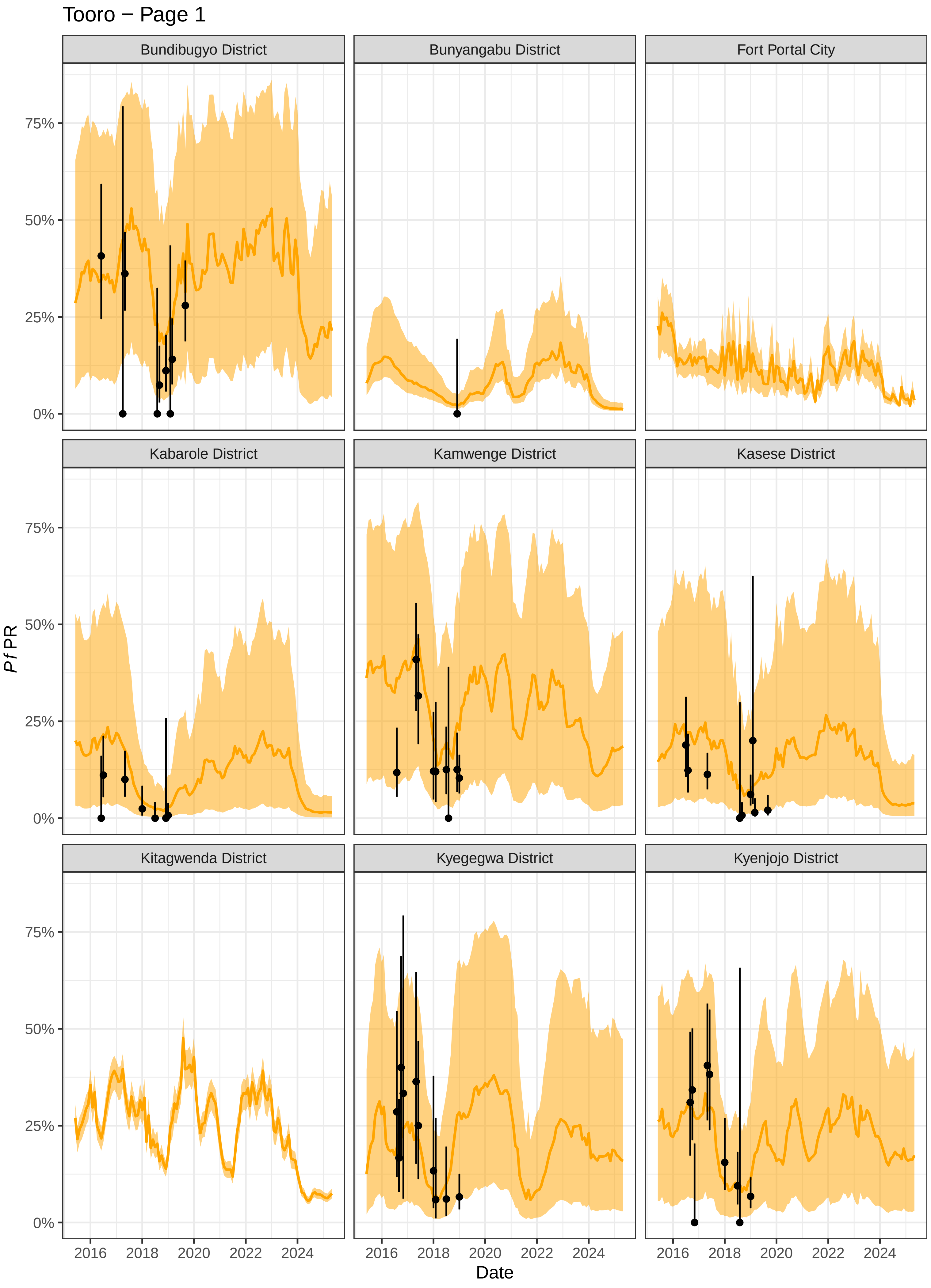
Tooro − Page 1
Bundibugyo District
Bunyangabu District
Fort Portal City
Kabarole District
Kamwenge District
Kasese District
Kitagwenda District
Kyegegwa District
Kyenjojo District
0%
25%
50%
75%
2016
2018
2020
2022
2024
Date
Pf PR

### Tooro − Page 2

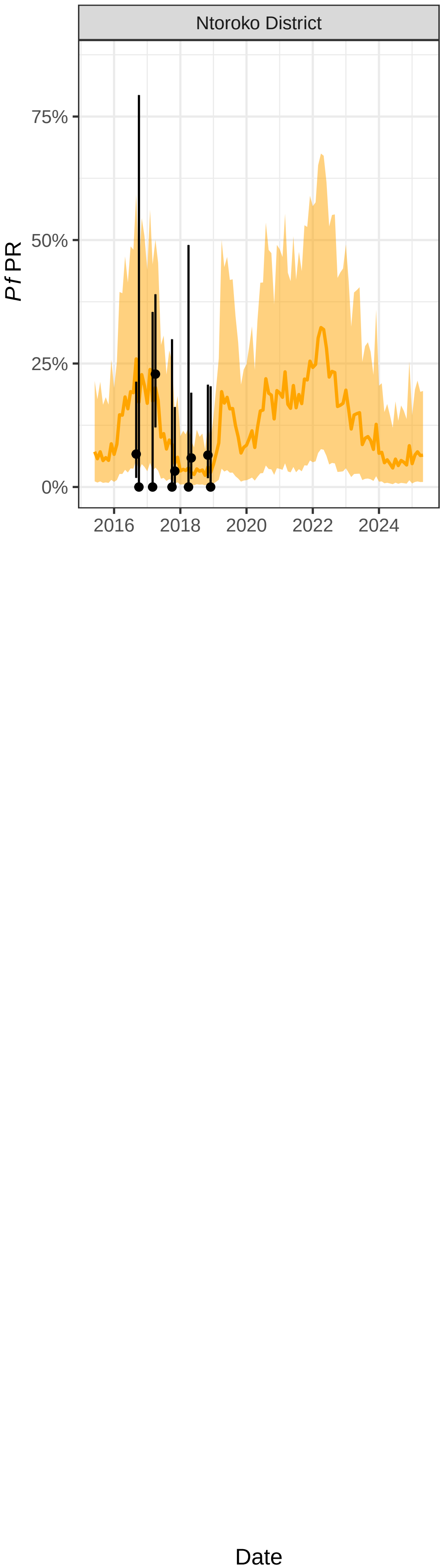

Ntoroko District
75%
50%
25%
0%
*Pf* PR
2016
2018
2020
2022
2024
Date

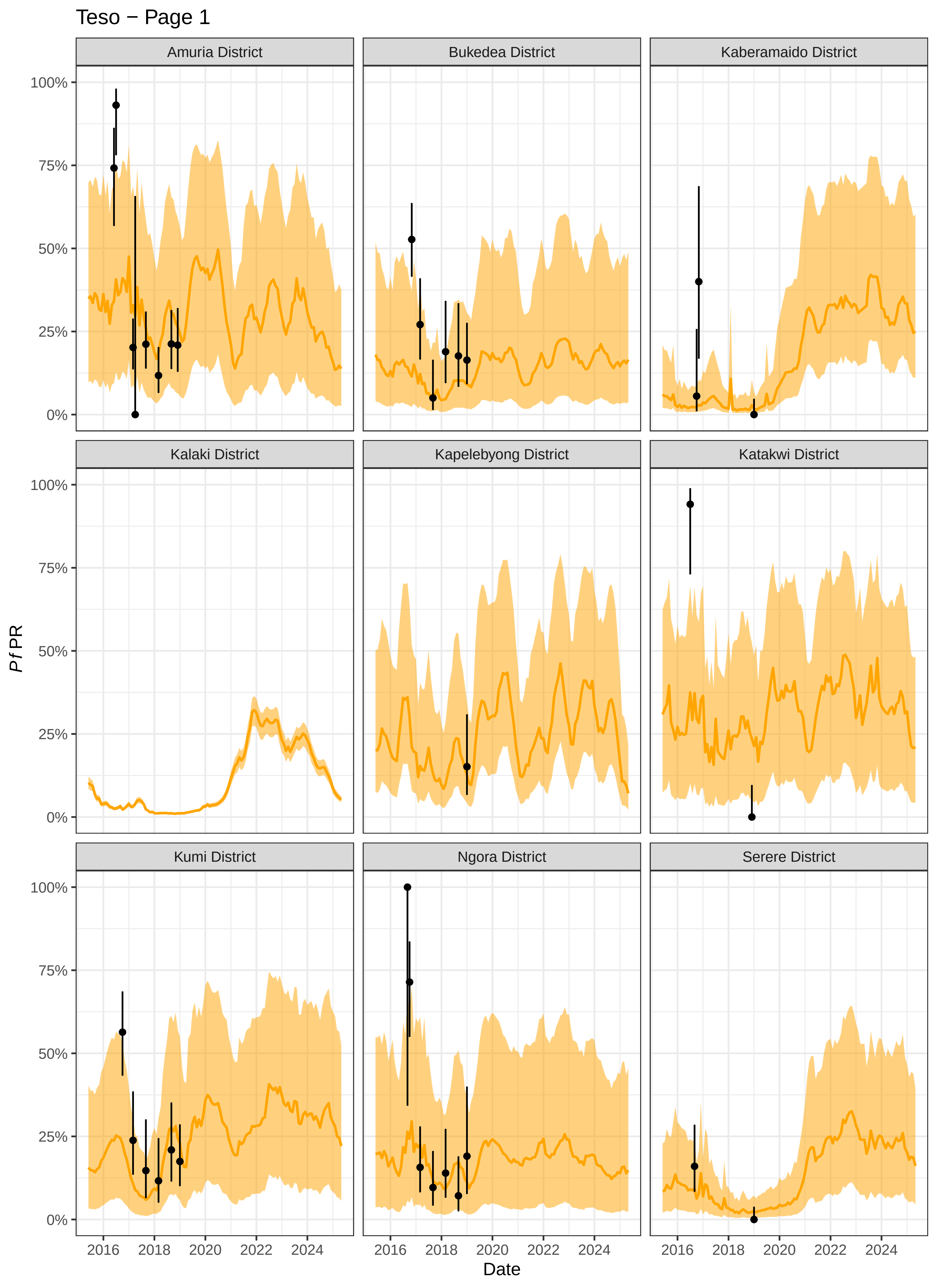

Teso − Page 1
Amuria District
Bukedea District
Kaberamaido District
Kalaki District
Kapelebyong District
Katakwi District
Kumi District
Ngora District
Serere District
100%
75%
50%
25%
0%
*Pf*PR
2016
2018
2020
2022
2024
Date

### Teso − Page 2

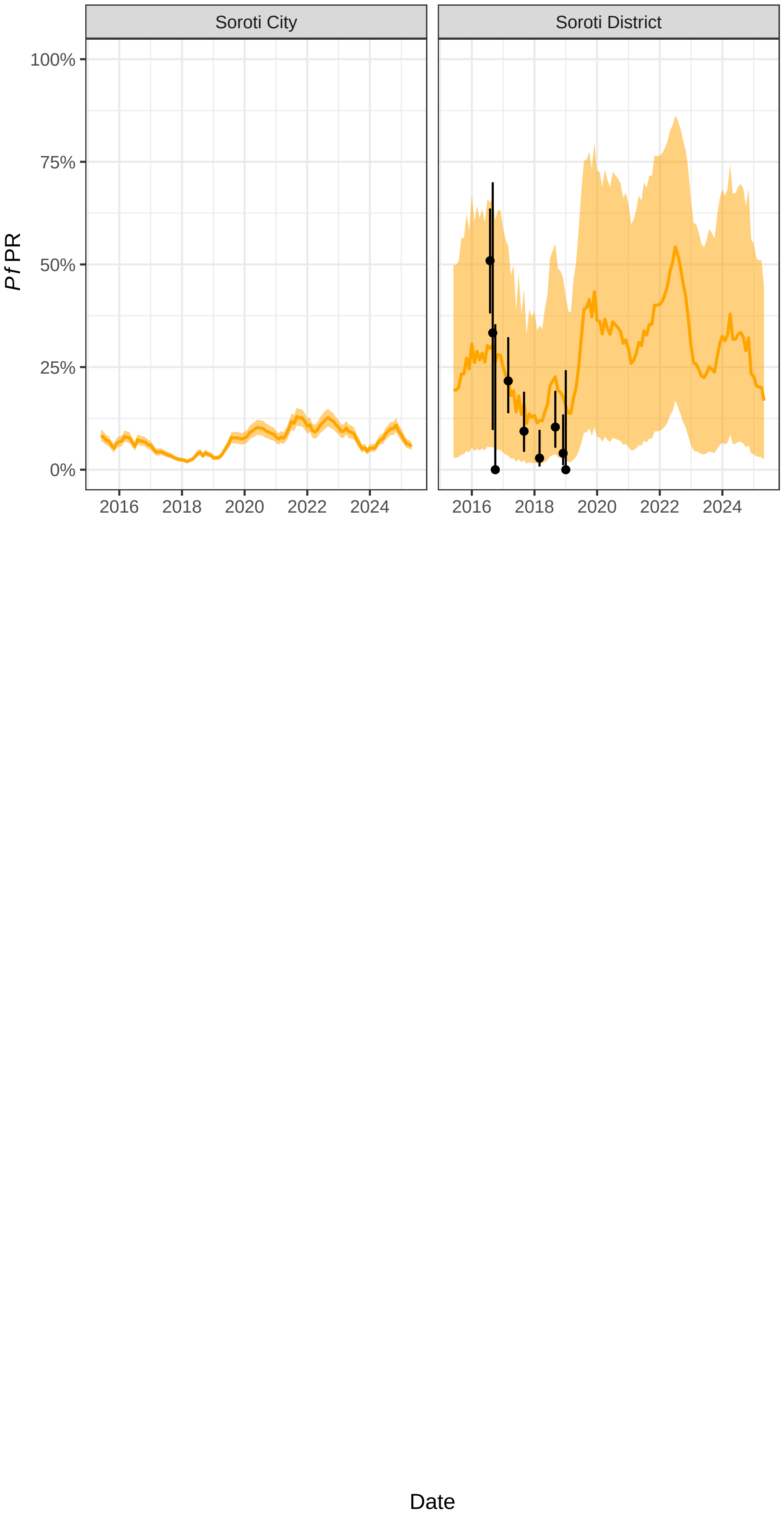
Soroti City
Soroti District
100%
75%
50%
25%
0%
Pf PR
2016
2018
2020
2022
2024
Date

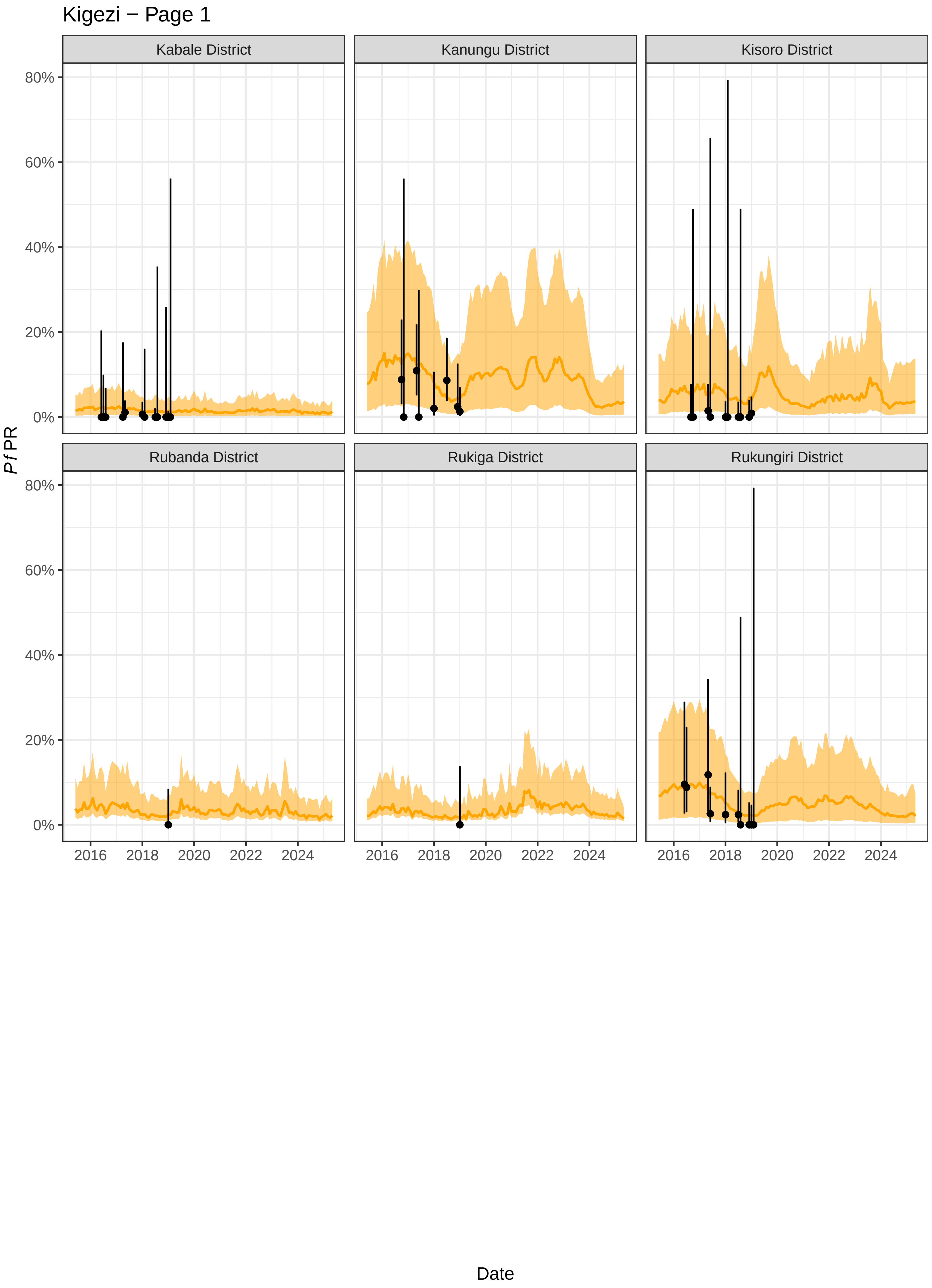

Kigezi − Page 1
Kabale District
Kanungu District
Kisoro District
Rubanda District
Rukiga District
Rukungiri District
80%
60%
40%
20%
0%
Pf PR
2016
2018
2020
2022
2024
Date

### Kampala − Page 1

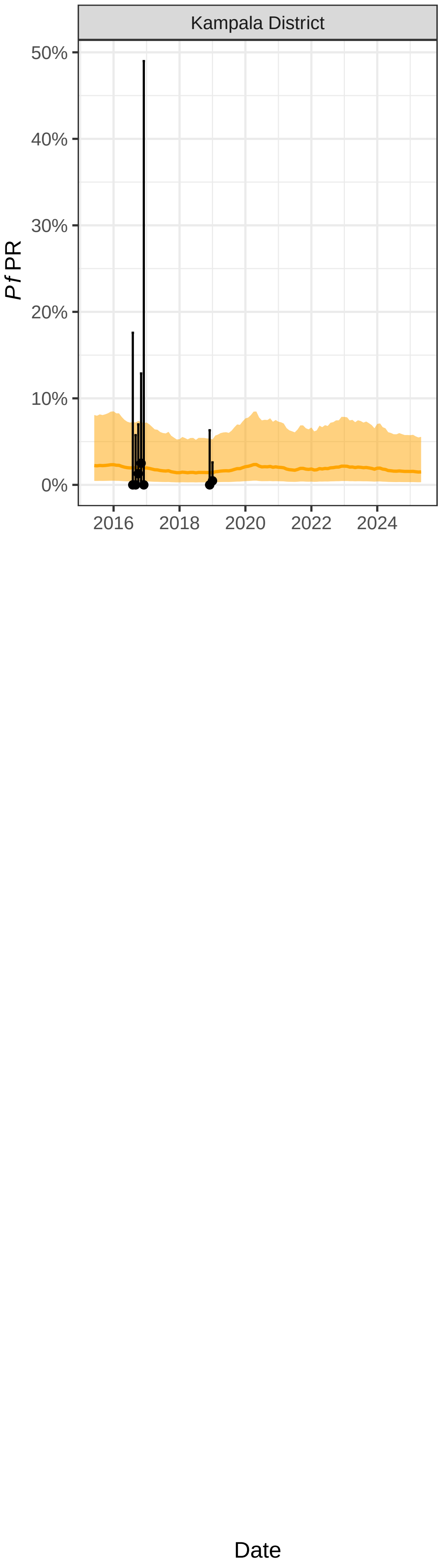
